# Sex-Specific Genetic Architecture and Comorbidities of Alcohol Use Behaviors

**DOI:** 10.64898/2025.12.01.25341089

**Authors:** Laura Vilar-Ribó, Mariela V Jennings, Aisha Sallah, Maria Niarchou, Zeal Jinwala, Hayley HA Thorpe, Sevim B Bianchi, John Meredith, Kyra Feuer, Lydia Rader, Natasia Courchesne-Krak, Jared Balbona, 23andMe Research Institute, Sarah L Elson, Pierre Fontanillas, Emma C Johnson, Lea K Davis, Alexander S Hatoum, Travis T Mallard, Daniel E Gustavson, Hang Zhou, Abraham A Palmer, Jeanne E Savage, Rachel L Kember, Sandra Sanchez-Roige

**Author notes:** **Corresponding Author:** Sandra Sanchez-Roige, PhD, Department of Psychiatry, University of California San Diego, 9500 Gilman Drive, La Jolla, CA, USA, 92093.

## Abstract

**Background:** Sex differences in alcohol use behaviors are well-established: males typically engage in heavier and more frequent drinking and exhibit more externalizing behaviors (e.g., other substance use), while females often transition to dependence more rapidly and present more internalizing psychopathology (e.g., depression). The biological mechanisms underpinning these differences are relatively unknown.

**Methods:** In this study, we investigated the sex-differentiated genetic architecture of 11 alcohol use phenotypes pertaining to frequency, quantity and problematic use by leveraging sex-stratified genome-wide association studies (Ns 40,335 to 613,148). Specifically, we compared SNP-based heritability (***h^2^_SNP_***) estimates, individual genetic locus effects, genetic correlations (***r_g_***) across alcohol phenotypes and with comorbid traits from independent GWAS, and polygenic score (**PGS**) associations with medical outcomes from clinical populations.

**Results:** *h^2^_SNP_* was broadly similar between sexes, except for higher estimates in males for *beer quantity* and *problematic alcohol use* (**PAU**). We identified four sex-differentiated top loci (*p_sex-diff_* < 5 x 10^-8^), including a female-specific association in *IZUMO1* for *drinking frequency* and *quantity*, and three male-specific associations in *ADH1B*, *KLB* and *FTO* for *beer quantity* and/or *PAU*. Between-sex genetic correlations ranged from 0.68±0.07 to 0.89±0.04, these estimates were lowest for quantity measures and varied by beverage type, indicating partially distinct polygenic architecture. In males, we identified stronger positive genetic correlations with several externalizing traits (e.g., general addiction) compared to females. In females, we identified a specific positive genetic correlation with a single internalizing trait, self-harm. PGS analyses revealed sex-specific medical associations (e.g., bone/musculoskeletal conditions in females; hepatic/respiratory/infectious sequelae in males) that were obscured in sex-combined analyses; however, sex-specific PGS did not outperform combined-sex PGS for predicting alcohol use disorder diagnosis.

**Conclusions:** Sex-aware analyses of alcohol use behaviors can improve our understanding of the genetic etiology of alcohol use and related health outcomes, and future studies should consider cultural variation (e.g., drinking attitudes, social norms) in the relationship between behavior and genetics.

## INTRODUCTION

Alcohol use disorder (**AUD**) afflicts over 15 million Americans, costs the US economy $249 billion annually, and kills more than 3 million people a year globally^1,2^. AUD develops through a dynamic process, which begins with the initiation of use, continues with the escalation to hazardous drinking, and can lead to compulsive, harmful use that persists despite negative consequences^3^. AUD often manifests alongside other psychiatric and medical comorbidities, such as liver disease, cardiovascular conditions, pancreatitis, and metabolic disorders^4–6^.

Although males are more likely to be diagnosed with AUD than females, this gap is closing in western countries^7^. The trajectory of AUD development, however, is sex specific. While males are more prone to heavy drinking, females often transition to dependence more quickly^8^, show more susceptibility to cravings^9^ and relapse^10^, and experience aggravated adverse events to AUD medications^11^. Alcohol-related psychiatric comorbidities are also sex-specific. For example, males are more likely to exhibit externalizing disorders (e.g., other substance use disorders [**SUD**]), whereas females are more likely to manifest internalizing psychopathology (e.g., depression;^12,13^. Even at lower consumption levels^14,15^ females are disproportionately affected by medical comorbidities (e.g., liver disease, neurological damage, cardiovascular conditions)^16–19^. Many factors, both biological (e.g., hormones, body composition, metabolism) and environmental (e.g., cultural norms, social roles, beverage preferences), can shape these trajectories differently across sexes^7,20^.

Twin studies show that sex-specific AUD trajectories and comorbidities arise in part from genetic factors^7,17,21^, with some studies reporting slightly lower heritability estimates in females (e.g., 40-45% in females, 50-52% in males). A recent genome-wide association study (**GWAS**) identified potential sex-differentiated genetic variants (i.e., variants that are significantly different in the magnitude or direction of effect between sexes) associated with problematic alcohol use (**PAU**)^22^. However, this study included substantially fewer females than males (N_females_ = 143,198 vs. N_males_ = 496,548), resulting in power imbalances between sexes. In addition, the genetic architecture of AUD is not identical to that of alcohol consumption^3,23–25^ and the sex-differentiated genetic architecture across the broader spectrum of alcohol use behaviors has yet to be explored. Identifying sex-differentiated genes and biological pathways underlying alcohol use behaviors and comorbidities may enable personalized AUD treatments and prevention strategies that are tailored to sex and accommodate heterogeneous profiles.

In the present study, we performed a systematic genomic characterization of sex differences across the spectrum of alcohol use behaviors. We leveraged data from previously published sex-stratified GWAS including PAU^22^, 9 alcohol use behavior phenotypes from UK Biobank^26^ (N_females_ = 67,076 - 191,917 vs. N_males_ = 53,332 - 168,772), and a new, unpublished sex-stratified GWAS of Alcohol Use Disorder Identification Test (**AUDIT**) total scores from 23andMe Research Institute (N_females_ = 72,444 vs. N_males_ = 40,335). We examined the sex-differentiated genetic architecture of these phenotypes, across both *quantitative* effects (i.e., the magnitude of genetic influence by sex) and *qualitative* effects (i.e., specific genetic variants). Specifically, we compared SNP-based heritability (***h^2^_SNP_***) estimates, individual genetic locus effects, genetic correlations (***r_g_***) across alcohol phenotypes and with comorbid traits from independent GWAS, and PGS associations with medical outcomes from clinical populations. We had three specific hypotheses. First, that sex-differentiated genetic effects would be more pronounced in alcohol phenotypes influenced by body size (e.g., body mass index [**BMI**]) and metabolic processes, such as drinking quantity, given that these traits tend to differ between females and males^27^. We expected that these differences would be less evident for phenotypes that reflect behavioral or psychiatric components of AUD, such as drinking frequency or problematic alcohol use. Second, that externalizing comorbidities would be more strongly associated in males, whereas internalizing comorbidities would show stronger associations in females based on epidemiologic and twin evidence of sex-differentiated comorbidity profiles^28,29^. Third, although we anticipated that sex-combined PGS, which are trained on pooled datasets with larger sample sizes, would outperform sex-specific PGS in predicting AUD diagnosis^30–33^, we hypothesized that sex-specific PGS would uncover associations with sex-specific comorbidities that may be masked in sex-agnostic analyses.

## METHODS

### Sex-stratified GWAS

We included sex-stratified GWAS summary statistics for 11 alcohol use phenotypes across three categories: frequency (*drinking frequency, drinking with meals* and *decreased drinking*), quantity (*drinking quantity, beer quantity, spirit quantity, red wine quantity* and *white wine quantity*) and problematic use (*binge drinking, AUDIT total score* and *PAU*) (**Table 1**).

**Table 1.**
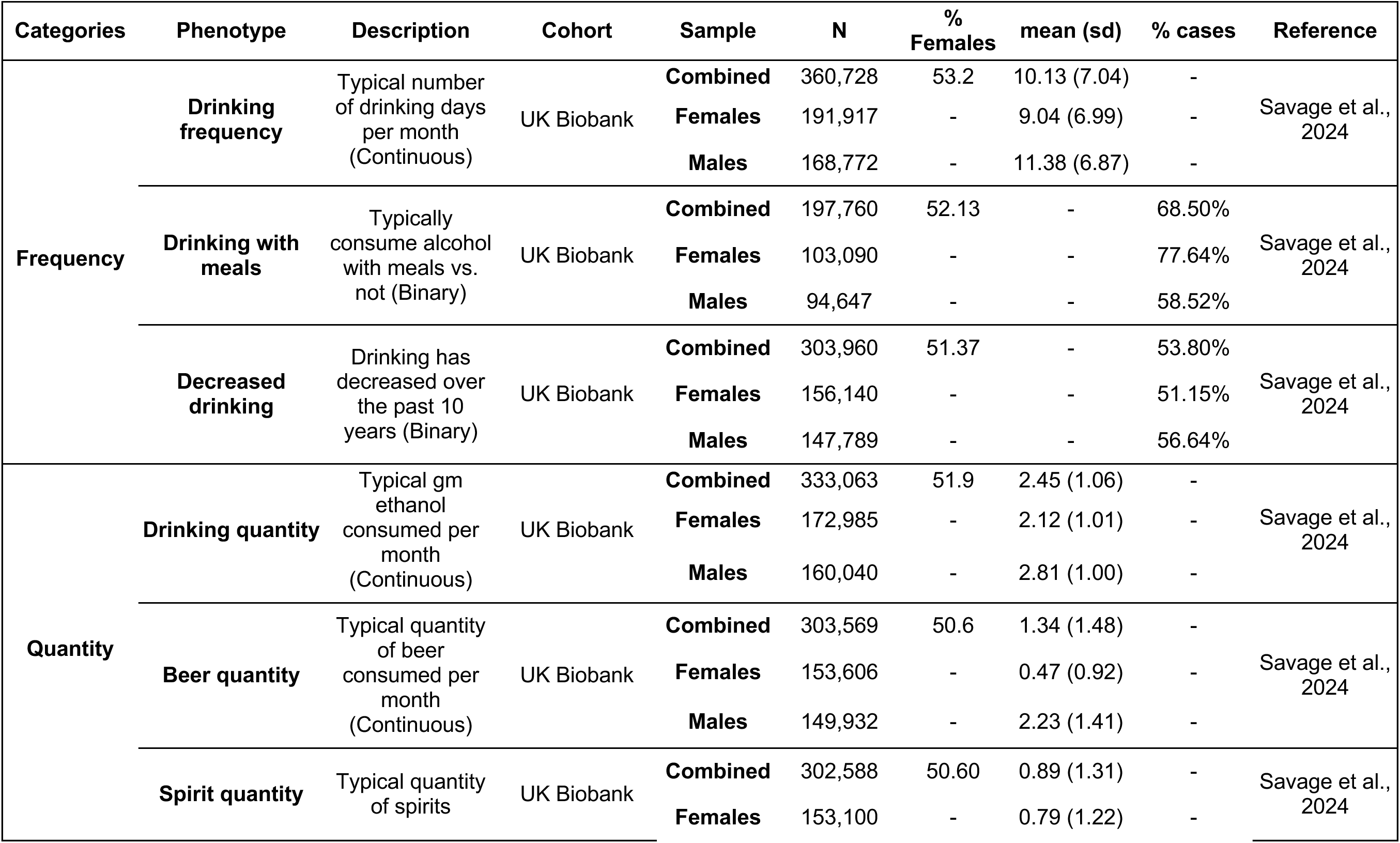
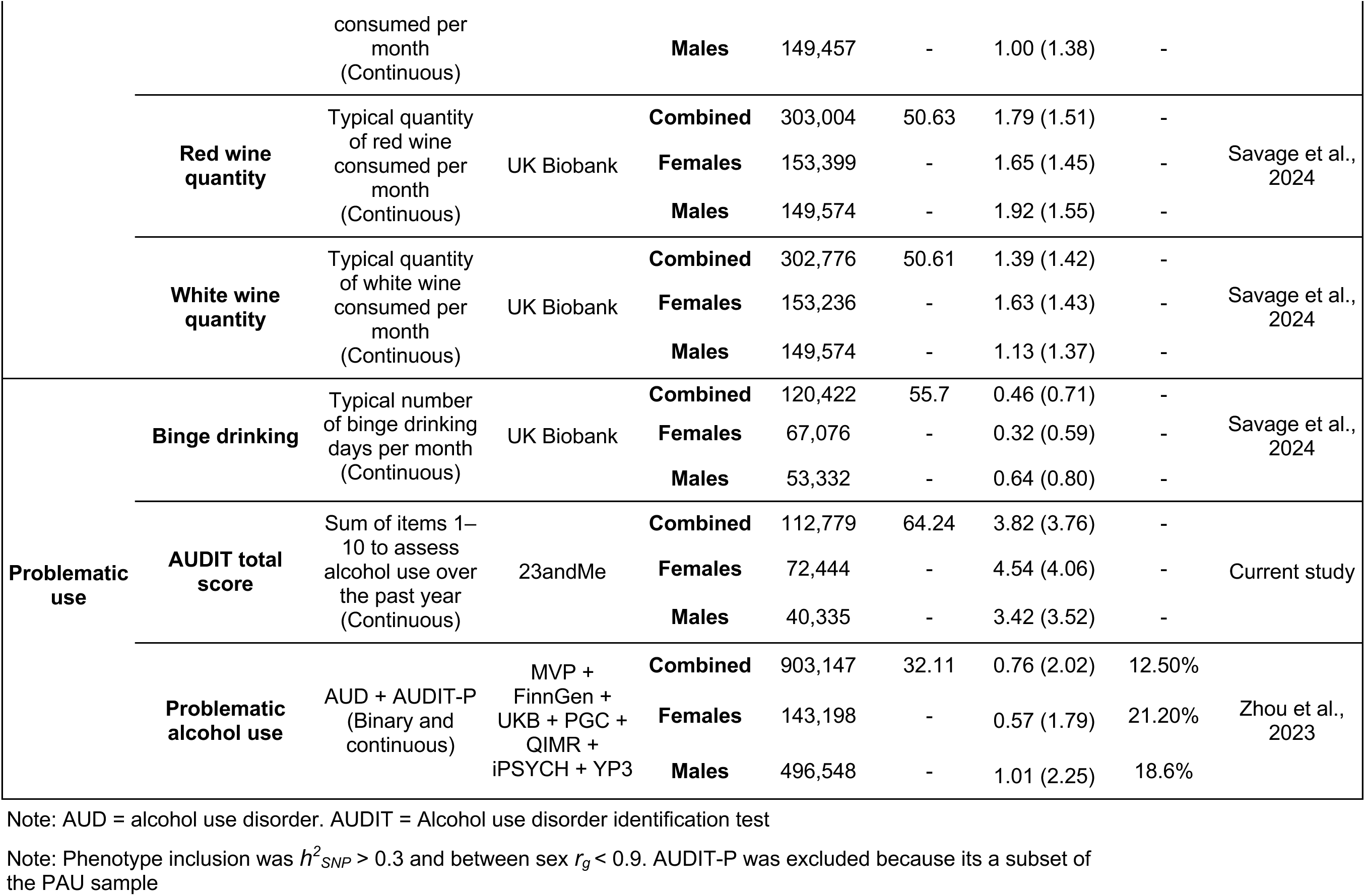
Summary of the 11 alcohol use phenotypes included in the study, spanning measures of frequency, quantity, and problematic use. Results are presented for sex-combined as well as female- and male-specific GWAS datasets. For continuous phenotypes, mean values and standard errors are reported, while for binary phenotypes the proportion of cases is provided.

Phenotypes were selected based on prior evidence of differences in between-sex genetic correlation (*r_g_* < 0.9)^26^. We additionally excluded phenotypes with non-significant or low *h^2^_SNP_* in the sex-combined sample (*h^2^_SNP_* < 0.03 and/or Z-score < 10) to ensure reliable results^34^. The full list of phenotypes excluded based on these criteria is shown in the **Supplemental Material** and in **Figure S1**. All GWAS were restricted to individuals of genetically inferred European ancestry based on reference populations due to the unavailability of adequately powered sex-stratified GWAS in other ancestral populations^35,36^. Methods used to perform the sex-stratified AUDIT total score GWAS can be found in the **Supplemental Material**. Methods used to perform the sex-stratified GWAS on the remaining datasets can be found in the original publications^22,26^.

### Sex-specific heritability and genetic correlation analyses

We calculated *h^2^_SNP_* using linkage disequilibrium score regression (**LDSC**)^37^. SNPs were mapped to HapMap3 data using pre-computed linkage disequilibrium scores included with the software (“eur_w_ld_chr”). For binary phenotypes, we estimated *h^2^_SNP_* on the liability scale using population prevalences (**Table S1**). We also used LDSC to estimate genetic correlations (*r_g_*) among alcohol use phenotypes, both between and within sexes, as well as with 71 complex traits from published sex-combined GWAS related to alcohol use and AUD based on prior literature, including other substance use, externalizing, internalizing, cognitive, sociodemographic and anthropometric traits^5,38–40^. For each alcohol phenotype, we applied a 5% FDR threshold to account for multiple testing.

### Sex-differentiated genetic effects

To identify loci with sex-differentiated genetic effects, we compared SNP effect sizes derived from sex-stratified GWAS of each alcohol use phenotype by calculating Z-scores and corresponding *p*-values across the genome. We focused on loci with genome-wide significant sex-differentiated effects (*p_sex-diff_* < 5 x 10^-8^) and that reached genome-wide significance in one sex but not in the other. While the most direct approach to identify SNPs with sex-differentiated effects is through genotype-by-sex interaction analyses, it requires access to individual-level data. A widely used alternative, which is statistically similar when covariate interactions and variance differences are equivalent in the two sexes, is to compare effect sizes from sex-stratified GWAS using a Z-score-based difference test^41^.

### Polygenic score phenome-wide association analyses (PheWAS)

To investigate sex-specific medical associations, we conducted PheWAS of both sex-specific and sex-combined alcohol use PGS against thousands of medical outcomes in two clinical cohorts (described below). PheWAS were meta-analyzed across sites using fixed effects in METAL v0.99.6.1^42^. We examined the association between sex-specific PGS in sex-stratified samples and sex-combined PGS in the total sample for each alcohol use phenotype. Medical outcomes were derived by mapping ICD-9 and ICD-10 codes to 1,817 “phecodes”, as described and validated by the Phecode Map 1.2b^43^. Phecodes use a standardized vocabulary and denote diseases, traits, or symptoms (http://phewascatalog.org). We excluded phecodes with less than 100 cases and phecodes that were sex-specific (i.e., unique to one sex, such as endometriosis).

#### Vanderbilt University Medical Center biobank (BioVU)

The BioVU cohort was composed of 66,914 unrelated individuals (females = 37,239; males = 29,675) who provided electronic health records (**EHR**) to the Vanderbilt University Medical Center (VUMC; IRB #160302, #172020, #190418)^44^. Genotyping used the Illumina MEGA_ex platform, with imputation via the Michigan Imputation Server^45^. SNPs with imputation quality (*R^2^* > 0.3) were converted to hard genotype calls. We applied an IBD filter of 0.2 to exclude cryptic relatedness. Genetic ancestral groups were determined based on principal component analysis (**PCA**) and comparison to known ancestries in the 1000 Genomes Project Phase 3^46^ reference panel; we restricted the analyses to individuals most genetically similar to European reference populations. We also applied a minor allele frequency (**MAF**) filter of 0.05. Details on genotyping and quality control are available for VUMC^47^.

#### Penn Medicine BioBank (PMBB)

PMBB is an EHR-linked biobank at the University of Pennsylvania (Penn Medicine) with 44,297 participants. PMBB samples were genotyped by the GSA genotyping array. Quality control removed SNPs with marker call rate <95% and sample call rate <90%, and individuals with sex discrepancies. Genotype phasing and imputation was performed on the TOPMed Imputation server^45^. The phasing was done using EAGLE (v2.4.1)^48^ and imputation was performed using MINIMAC software^45^. IBD analysis was used to check for relatedness among imputed samples using PLINK v.1.9^49^. We randomly removed one individual from each pair of related individuals (pihat <0.25). SNPs with an INFO score <0.3, MAF <0.01, a genotype call rate <0.95 or an HWE p<1.00E-6 were removed and restricted the analyses to individuals most genetically similar to European reference populations (N = 29,160; females = 12,998; males = 16,162). To estimate genetic ancestry, principal components (**PC**) were calculated based on common SNPs between PMBB and the 1000 Genomes Project phase 3^46^ using the smartpca module of the Eigensoft package^50^. Participants were assigned to an ancestry based on the distance of 10 PCs from the 1000 Genomes reference populations.

We computed PGS using PRS-Continuous shrinkage software (**PRS-CS**)^51^ using default “auto” settings to estimate shrinkage parameters and random seed fixed for reproducibility. Logistic models were fitted for each of the phecodes using the PheWAS v0.12 R package^43^ adjusting for sex when appropriate, median age of longitudinal EHR, and the first 10 PCs of genotype. To quantify the variance in AUD explained by each PGS, the adjusted partial Nagelkerke pseudo-*R²* was computed using the rsq.partial() function from the *rsq* R package v.2.7^52^. The weighted average Nagelkerke *R²* across sites was calculated with the formula Nagelkerke-*R²_WA_* = ∑ w_i_\**R*_i_*²*/∑ w_i_. For each alcohol phenotype, we used a 5% false discovery rate (**FDR**) threshold to correct for multiple testing within each phenotype.

### Statistical tests to estimate sex differences

Sex differences were quantified following best practices^41^ by computing a Z-score of the test statistics: Z = (X_F_ − X_M_)/sqrt(se_F_^2^ + se_M_^2^), where X represents *h^2^_SNP_*, SNP effect sizes, *r_g_* or odds ratio (OR) and se represents the standard error, for females and males, respectively. This test is the standard approach for assessing differences between independent parameters with known standard errors and under the assumption that the estimates are normally distributed, a condition typically met in GWAS due to large sample sizes^53^. A two-tailed test was performed to obtain the associated *p_sex-diff_* value. We applied a 5% FDR threshold to account for multiple testing for all tests, except for the sex-differentiated effect sizes, where genome-wide significance was defined as *p_sex-diff_* < 5 x 10^-8^.

## RESULTS

### SNP-based heritability

All 11 alcohol use phenotypes had significant *h^2^_SNP_* estimates, ranging from 0.031 ± 0.003 to 0.128 ± 0.007 (**Figure S2**; **Table S2**). Only two phenotypes showed statistically significantly greater *h^2^_SNP_* estimates in males compared to females: *beer quantity* (male *h^2^_SNP_ =* 0.097 ± 0.006, female *h^2^_SNP_* = 0.031 ± 0.003, *p_sex-diff_* = 3.83e-23) and *PAU* (male *h^2^_SNP_ =* 0.083 ± 0.004, female *h^2^_SNP_* = 0.045 ± 0.005, *p_sex-diff_* = 3.94e-09). The remaining alcohol use phenotypes showed similar *h^2^_SNP_* estimates across sexes (**Table S2**).

### Sex-differentiated genetic effects

**Figure 1A** shows the number of independent genome-wide significant loci in females (range = 1-23) and males (range = 0-40). While the magnitude of effect across variants was largely similar across sexes (**Figure S3 and S4**), we identified four independent loci with significant sex-differentiated effects (*p_sex-diff_* < 5 x 10^-8^) that reached genome-wide significance in one sex but not in the other across four alcohol traits (**Figure 1B**, **Table S3**). One locus (lead SNP rs838147 in *IZUMO1*) showed female-specific effects for both *drinking frequency* (OR_F_ = 0.87 [0.83, 0.91]) and *drinking quantity* (OR_F_ = 0.98 [0.97, 0.99]; **Figure S5A-B**). Two loci (lead SNPs rs1229984 in *ADH1B*, rs28712821 in *KLB)* showed male-specific effects for *beer quantity* (OR_M_ = 0.81 [0.77, 0.84] and OR_M_ = 0.96 [0.95, 0.97], respectively) and one locus (lead SNP rs1421085 in *FTO*) showed male-specific effects for *PAU* (OR_M_ = 1.02 [1.02, 1.03]; **Figure S5C-E**). Two of these loci (rs838147 and rs1421085) showed decreased risk in females but increased risk in males (**Figure 1C**). The other two loci (rs1229984 and rs28712821) showed decreased risk in both sexes, but with significantly larger magnitudes in males.

**Figure 1.**
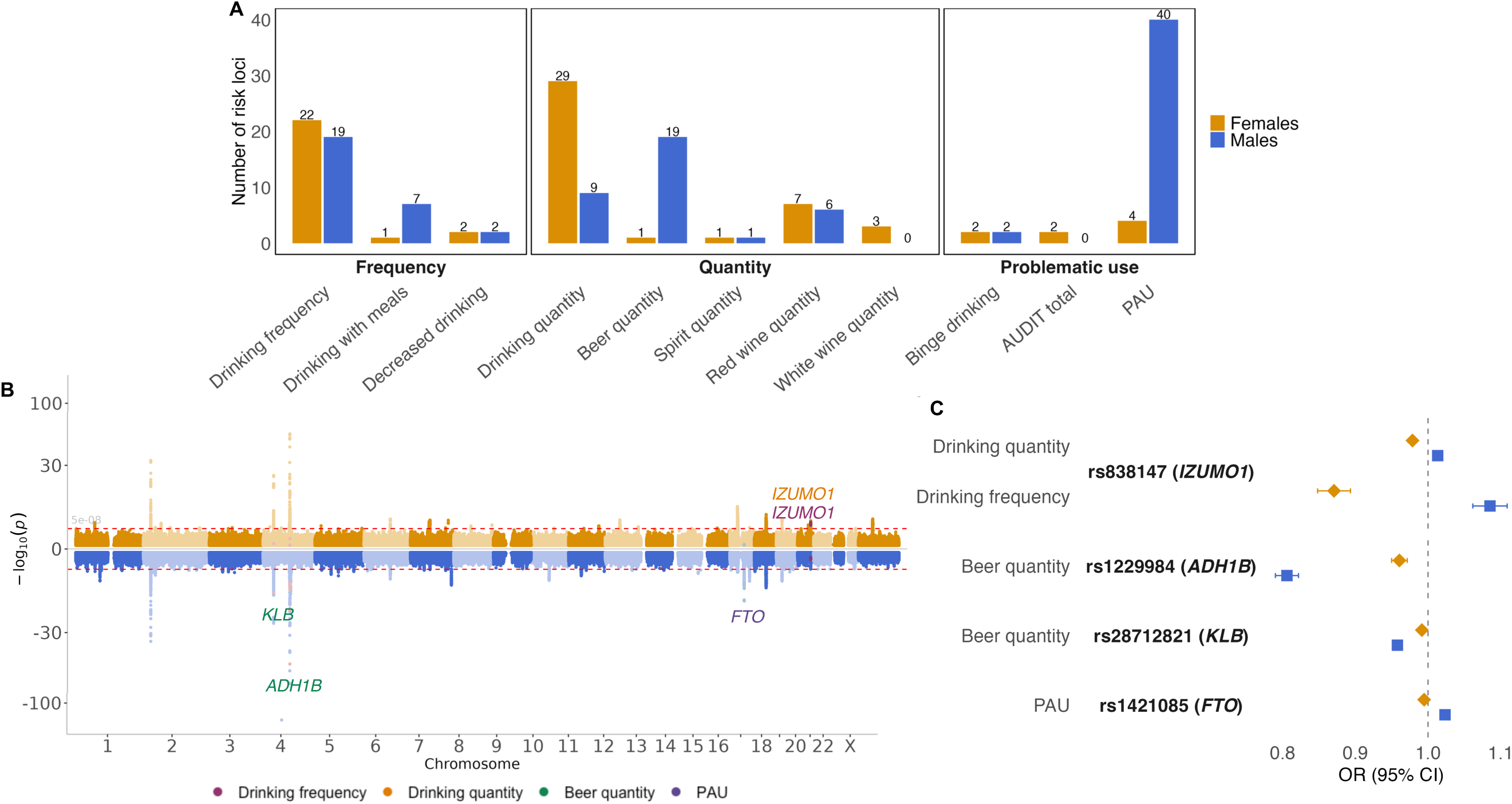
Number of genome-wide significant loci and loci with sex-differentiated genetic effects. **A.** Bar plot with the number of independent genome-wide significant loci for each alcohol use phenotype in females (amber) and in males (blue) **B.** Miami plot displaying the 11 sex-stratified GWAS overlaid on top of each other in females (top, in amber) and in males (bottom, in blue). Sex-differentiated loci reaching genome-wide significance are annotated with the nearest gene and colored by alcohol use phenotype: *drinking frequency* (dark purple), *drinking quantity* (turquoise), *beer quantity* (green), and *PAU* (violet). Miami plots for all alcohol use phenotypes can be found in **Figure S3. C.** Forest plot showing odds ratios (OR) and 95% confidence intervals for the lead SNPs for the four loci with sex-differentiated effects. ORs are shown separately for females (amber diamonds) and males (blue squares), illustrating the direction and magnitude of association in each sex. The nearest gene is shown in parentheses.

### Between-sex genetic correlations

Genetic correlations between sexes ranged from 0.68 ± 0.07 to 0.89 ± 0.04 (**Figure 2A**, **Table S4**). Consistent with our first hypothesis, drinking quantity phenotypes showed the lowest between-sex *r_g_* average (mean *r_g_* = 0.78 ± 0.06) compared to frequency (mean *r_g_* = 0.87 ± 0.06) or problematic use (mean *r_g_* = 0.85 ± 0.08). Of note, *r_g_* values across drinking quantity phenotypes varied widely depending on the beverage types (from 0.70 ± 0.06 for *beer quantity* to 0.89 ± 0.04 for *red wine quantity*).

**Figure 2.**
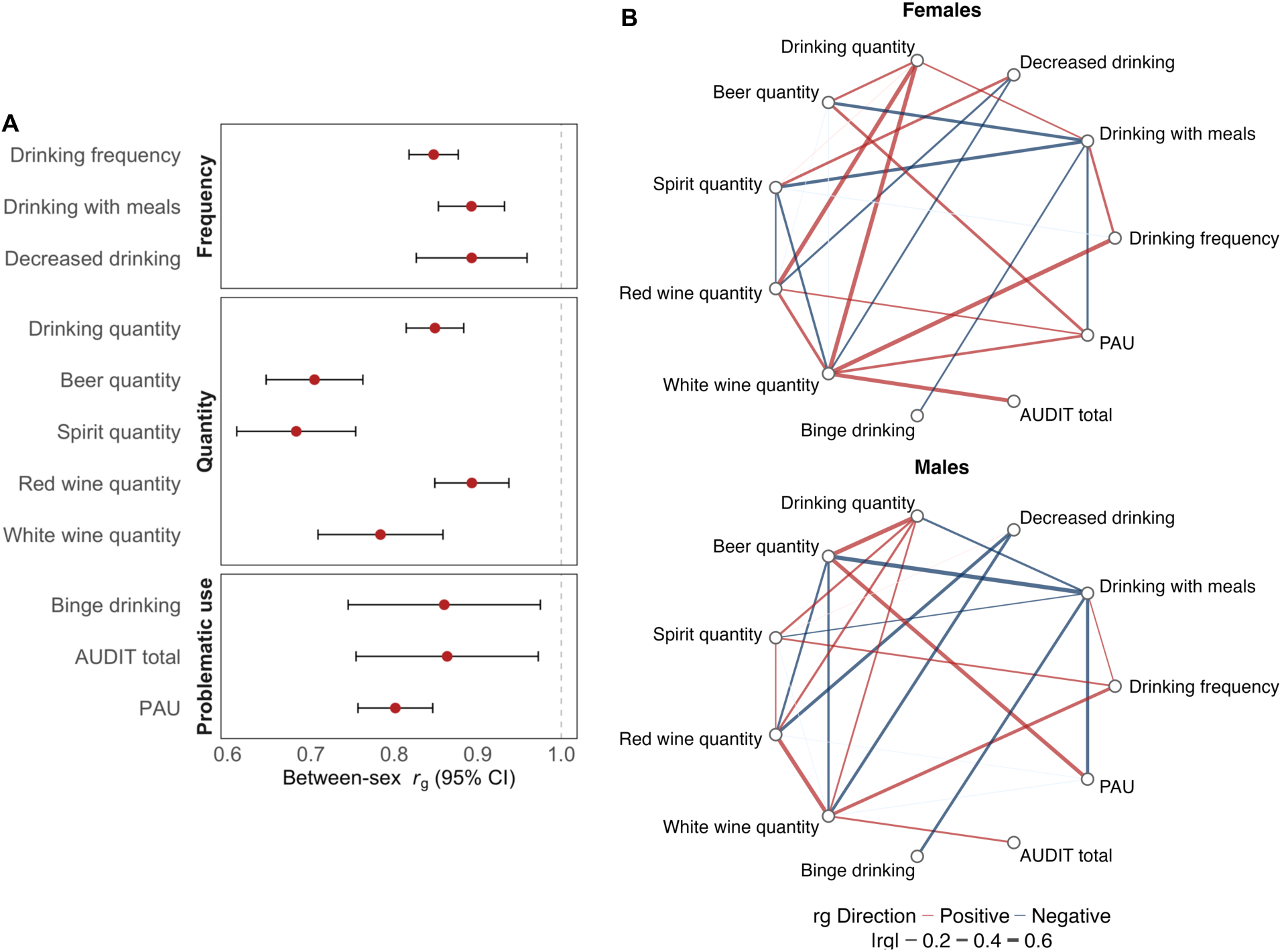
Genetic correlation (*r_g_*) estimates between sexes and among alcohol use phenotypes in females and males. **A.** Forest plot of between-sex *r_g_* with 95% CI across the 11 alcohol use phenotypes. Alcohol use phenotypes are grouped by category: frequency, quantity, and problematic use. **B.** Network plot of cross-trait genetic correlations in females (upper) and males (lower) for trait pairs with statistically significant sex differences in *r_g_* estimates. Edges represent pairwise genetic correlations: red edges indicate positive correlations, blue edges indicate negative correlations, and edge thickness reflects the absolute magnitude of *r_g_*. Faint (lighter) edges denote correlations that were not statistically significant within the respective sex. 10 pairs of traits showed stronger *r_g_* estimates in females and 12 pairs of traits showed stronger *r_g_* estimates in males.

### Genetic correlations among alcohol phenotypes

To test whether *r_g_* patterns across alcohol use phenotypes differed by sex, we computed cross-trait *r_g_* separately in males (*r_gM_*) and females (*r_gF_*). A total of 24 out of 55 (43.6%) trait-pair comparisons showed significant differences between sexes (**Figure 2B**, **Table S5**).

Thirteen significant pairs of traits showed consistent direction in *r_g_* estimates in females and males but were of larger magnitude in one of the sexes. The greatest differences in *r_g_* estimates were for *drinking quantit*y/*white wine quantity*, which were stronger in females (*r_gF_* = 0.68 vs. *r_gM_* = 0.23) and *drinking quantit*y/*beer quantity*, which were stronger in males (*r_gF_* = 0.30 vs. *r_gM_* = 0.72; **Figure 2B**, **Table S5**).

Two pairs of traits showed opposite direction in *r_g_* estimates by sex: *drinking quantit*y/*drinking with meals* were negatively genetically correlated in males but positively in females (*r_gF_* = 0.17 vs. *r_gM_* = -0.30), and *spirit quantity*/r*ed wine quantity* were positively genetically correlated in males but negatively in females (*r_gF_* = -0.14 vs. *r_gM_* = 0.13; **Figure 2B**, **Table S5**).

Nine pairs of traits showed significant *r_g_* estimates in only one sex. For example, *white wine quantity*/*PAU* were positively genetically correlated in females but not in males (*r_gF_* = 0.38 vs. *r_gM_* = -0.08) and *drinking quantity*/*spirit quantity* were positively genetically correlated in males but not in females (*r_gF_* = 0.07 vs. *r_gM_* = 0.31; **Figure 2B**, **Table S5**).

### Genetic correlations with other complex traits

We calculated *r_g_s* between alcohol use phenotypes in females and males and 71 other complex traits (**Table S6**). Of the *r_g_* estimates that were significant in either sex, 18.6% showed significant sex differences. Of these, 50.4% were concordant (i.e., same direction) across sexes, 6% were discordant (i.e., opposite directions), 21.7% were significant only in females (i.e., female-specific), and 21.7% were significant only in males (i.e., male-specific; **Figure 3A**). Quantity phenotypes showed greater sex differences in *r_g_* estimates compared to frequency or problematic use phenotypes (**Figure 3A**).

**Figure 3.**
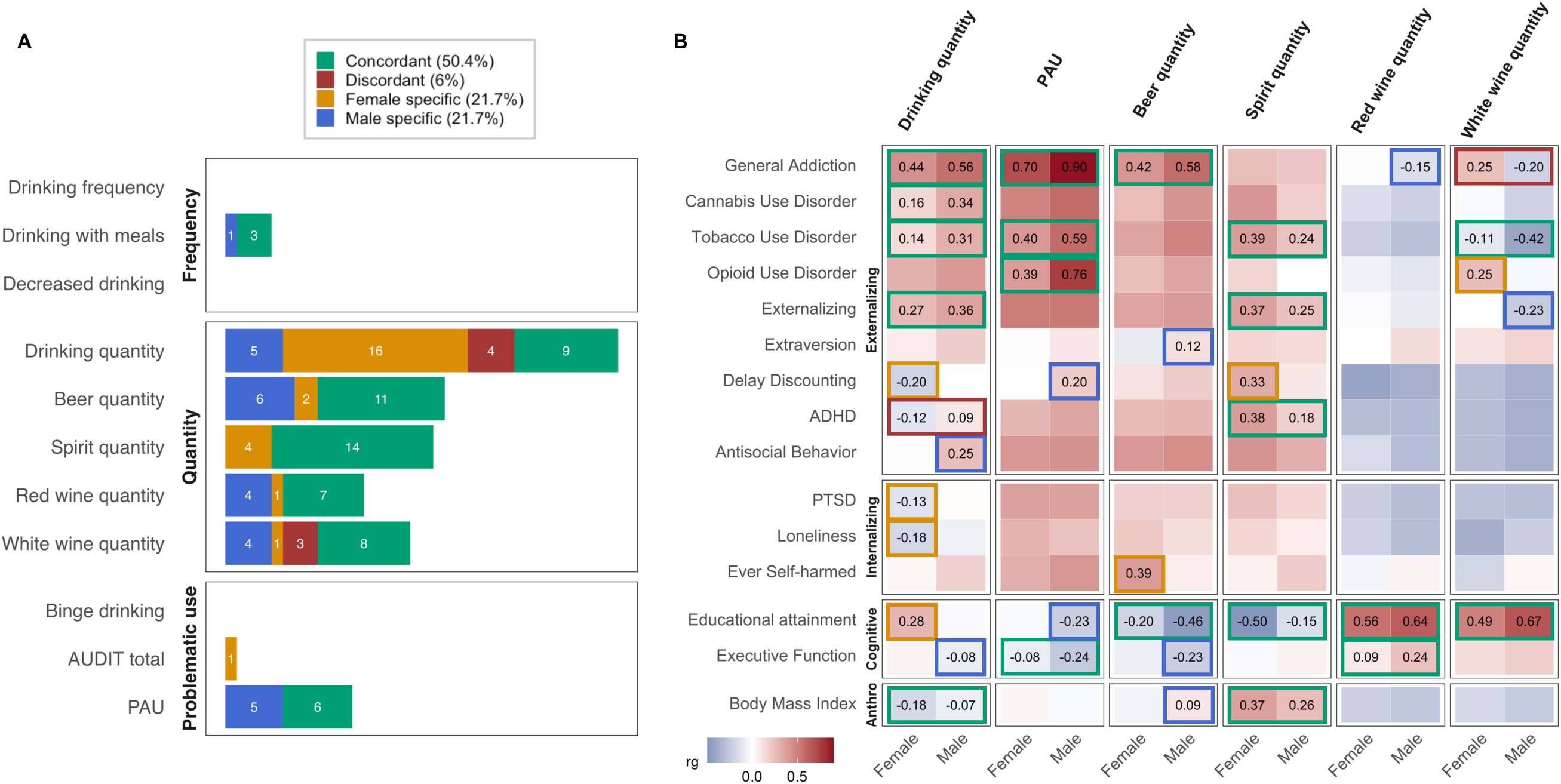
Genetic correlations (*r_g_*) between alcohol use phenotypes in females and males and 71 traits from other published sex-combined GWAS. **A.** Bar plot summarizing the number of traits showing significant sex differences in the *r_g_* estimates with each alcohol use phenotype. Results are categorized as: concordant (green) if significant in both sexes, with effects in the same direction; discordant (red) if significant in both sexes, but with effects in opposite directions; female-specific (amber) if significant only in females; and male-specific (blue) if significant only in males. No significant sex differences in *r_g_* were observed for *drinking frequency*, *decreased drinking* or *binge drinking*. **B**. Heatmap displaying *r_g_* estimates for 15 selected traits with significant sex differences (FDR corrected *p_sex-diff_* < 0.05). Traits are grouped into four categories: externalizing, internalizing, cognitive and anthropometric (Anthro). Only values for statistically significant *r_g_* estimates in each sex are shown. Cell color indicates the magnitude and direction of the correlation, ranging from blue (negative *r_g_*) to white (null) to red (positive *r_g_*). Cell outline indicates concordance, discordance, or sex-specificity as in panel A. ADHD: attention-deficit/hyperactivity disorder; PTSD: post-traumatic stress disorder.

**Figure 3B** shows a heatmap of *r_g_* for selected traits with significant sex differences (FDR corrected *p_sex-diff_* < 0.05). We observed similar *r_g_* patterns for overall *drinking quantity* and *PAU*; but identified greater variability in *r_g_* patterns when examining specific beverage types.

#### Substance use and externalizing traits

Our results were consistent with our second hypothesis, showing that *r_g_s* with externalizing traits are greater in males than in females. Among males we observed consistently stronger positive *r_g_s* between alcohol phenotypes and substance use traits compared to females. For example, *drinking quantity* was more strongly positively genetically correlated in males with general addiction (a genetically derived trait capturing shared liability to SUDs; *r_gF_* = 0.44 vs. *r_gM_* = 0.56), tobacco use disorder (*r_gF_* = 0.14 vs. *r_gM_* = 0.31), and cannabis use disorder (*r_gF_* = 0.16 vs. *r_gM_* = 0.34), and *PAU* was more strongly positively genetically correlated in males with tobacco use disorder (*r_gF_* = 0.40 vs. *r_gM_* = 0.59) and opioid use disorder (*r_gF_* = 0.39 vs. *r_gM_* = 0.76). The *r_g_s* with externalizing traits were also stronger in males. For example, *drinking quantity* was more strongly positively genetically correlated in males with externalizing psychopathology (*r_gF_* = 0.27 vs. *r_gM_* = 0.36), attention-deficit/hyperactivity disorder (**ADHD**; *r_gM_* = 0.09 ± 0.03 vs. -0.12 ± 0.03, *p* = 8.41e-05), and antisocial behavior (*r_gF_* = -0.12 vs. *r_gM_* = 0.25), and *PAU* was more strongly positively genetically correlated in males with delay discounting (*r_gF_* = -0.1 vs. *r_gM_* = 0.20). In contrast, we observed sex-specific *negative r_g_s* with externalizing traits for *drinking quantity* (e.g., delay discounting *r_gF_* = -0.20 vs. *r_gM_* = 0.001) in females.

For specific beverages, *beer quantity* was the only beverage in males that showed positive *r_g_s* with externalizing (e.g., general addiction (*r_gF_* = 0.42 vs. *r_gM_* = 0.58) and extraversion (*r_gF_* = -0.9 vs. *r_gM_* = 0.12)). On the contrary, *red wine quantity* (e.g., general addiction *r_gF_* = 0.01 vs. *r_gM_* = - 0.15) and *white wine quantity* showed negative *r_g_s* with externalizing traits in males (e.g., general addiction *r_gF_* = 0.25 vs. *r_gM_* = -0.2). In contrast, all *r_g_s* with externalizing traits were positive in females, particularly for *spirit quantity* (e.g., tobacco use disorder *r_gF_* = 0.39 vs. *r_gM_* = 0.24) and for *white wine quantity* (e.g. general addiction *r_gF_* = 0.25 vs. *r_gM_* = -0.20).

#### Internalizing traits

Results for internalizing traits were broadly consistent with our second hypothesis of stronger associations in females, but the evidence was modest. We observed female-specific *r_g_s* across only three traits. For *drinking quantity,* we observed female-specific negative *r_g_s* with post-traumatic stress disorder (PTSD; *r_gF_* = -0.13 vs. *r_gM_* 0.01) and loneliness (*r_gF_* = -0.18 vs. *r_gM_* = -0.06). However, for specific beverage types, particularly for *beer quantity,* we observed a female-specific positive *r_g_* with ever self-harmed (*r_gF_* = 0.39 vs. *r_gM_* = 0.07).

#### Cognitive traits

For cognitive traits, *r_g_* patterns were divergent across sexes for *drinking quantity* and *PAU*. We observed male-specific negative *r_g_s* with cognitive traits for *drinking quantity* (e.g., executive function *r_gF_* = 0.05 vs. *r_gM_* = -0.08) and for *PAU* (e.g., educational attainment *r_gF_* = -0.03 vs. *r_gM_* = - 0.23). In contrast, we observed female-specific positive *r_g_s* with cognitive and socioeconomic traits for *drinking quantity* (e.g., educational attainment *r_gF_* = 0.28 vs. *r_gM_* = -0.03).

Sex differences in *r_g_* patterns across cognitive traits varied across beverage types. In males, we observed stronger negative *r_g_* for *beer quantity* and educational attainment (*r_gF_* = -0.20 vs. *r_gM_* = -0.46) and executive function (*r_gF_* = -0.07 vs. *r_gM_* = -0.23), but stronger positive *r_g_s* for *red wine quantity* and executive function (*r_gF_* = 0.09 vs. *r_gM_* = 0.24) and for *white wine quantity* and educational attainment (*r_gF_* = 0.49 vs. *r_gM_* = 0.67). In contrast, in females, we observed stronger negative *r_g_s* for *spirit quantity* and educational attainment (*r_gF_* = -0.50 vs. *r_gM_* = -0.15).

#### Anthropometric traits

In females, we observed a stronger negative *r_g_* for *drinking quantity* and BMI than in males (*r_gF_* = -0.18 vs. *r_gM_* = -0.07). However, when looking at the *r_g_* for specific beverage types, the *r_g_* with BMI became positive for *spirit quantity* in females (*r_gF_* = 0.37 vs. *r_gM_* = 0.26), and for *beer quantity* in males (*r_gF_* = -0.05 vs. *r_gM_* = 0.09).

### Phenome-wide association studies

To test our third hypothesis that sex-specific PGS would uncover associations with sex-specific comorbidities that may be masked in sex-agnostic analyses, we investigated sex differences in comorbidity patterns between alcohol PGSs and other medical outcomes (**Table S7**). Specifically, we tested female PGSs in the female sample, and male PGSs in the male sample and benchmarked the results against the sex-combined PGSs in the full sample. Out of the 1,112 traits tested, up to 487 traits were significantly associated with at least one alcohol use PGS in at least one sex. Of these, 97 (19.9%) PGS-trait comparisons showed statistically significant sex differences (**Figure 4A**): 41 (8.42%) were significant in females as well as in the sex-combined samples (i.e., female specific); 15 (3.1%) were significant only in females (i.e., female unique); 32 (6.6%) were significant in males as well as in the sex-combined samples (i.e., male specific); 6 (1.2%) were significant only in males (i.e., male unique); and 5 (1%) were concordant across sexes but showed stronger associations in males. No concordant trait associations were stronger in females. **Figure 4B** shows the PGS associations for selected traits with significant sex differences (FDR corrected *p_sex-diff_* < 0.05).

**Figure 4.**
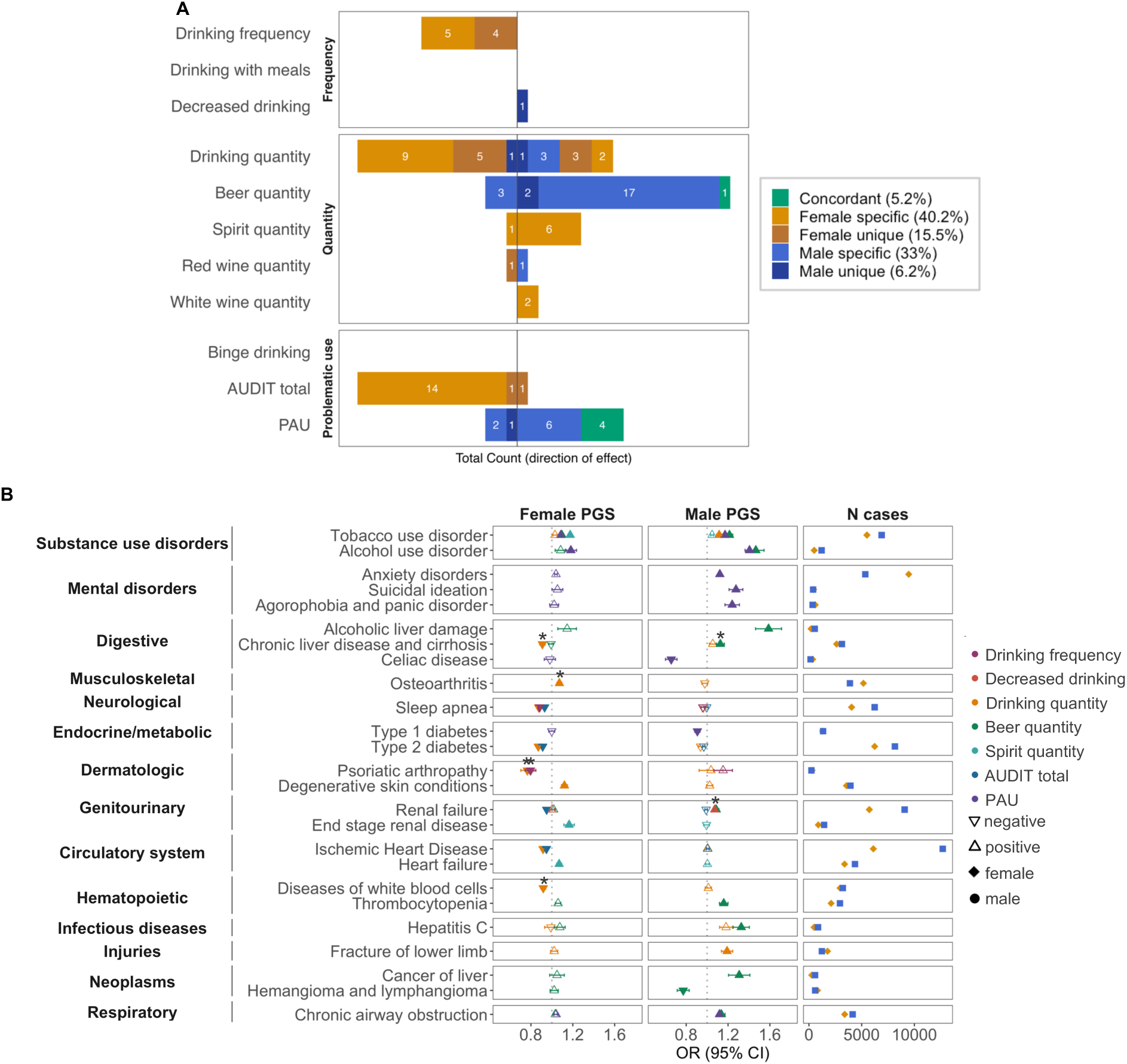
Phenome-wide association study (PheWAS) meta-analysis in BioVU and PMBB. **A.** Bar plot summarizing the number of traits showing significant sex differences between the female-specific and the male-specific polygenic scores (PGSs) associations across all alcohol use phenotypes. Results are categorized as: female-specific (amber) if significant in the female-specific and sex-combined PGS analyses; female-unique (dark amber) if significant only in the female-specific PGS analyses; male-specific (blue) if significant in the male-specific and sex-combined PGS analyses; male-unique (dark blue) if significant only in the male-specific PGS analyses; and concordant (green) if significant in both sexes, but with stronger effects in males. *Drinking quantity* and *AUDIT total* PGSs exhibited the most female-specific associations (15 and 14, respectively), while *beer quantity* and *PAU* exhibited the most male-specific associations (19 and 6, respectively). No significant sex differences in *r_g_* were observed for *drinking with meals* and *binge drinking*. **B.** Associations between alcohol use phenotypes PGSs and selected medical conditions across disease categories. PheWAS results are displayed for female PGS in the female target sample (left), and male PGS in the male target sample (middle) PGSs. Odds ratios (OR) are shown for selected phenotypes exhibiting sex-differentiated effects. Triangle direction indicates the direction of the association (upward for OR > 1, downward for OR < 1), and triangle color corresponds to the alcohol use phenotype used to construct the PGS. Filled triangles represent FDR-corrected significant associations, while empty triangles indicate non-significant associations. Asterisks mark female and male PGS associations that were not significant in the sex-combined PGS analyses (i.e., female-unique and male-unique associations). The right panel displays the number of cases in each medical condition for females (amber diamonds) and males (blue squares).

In the female PGS analyses, 14 (26%) of the sex-specific associations were positive (i.e., linked to increased risk for medical conditions), particularly for *drinking quantity* (n = 5) and *spirit quantity* (n = 6) PGSs (**Figure 4A**). These included associations with higher risk for other SUDs (e.g., tobacco use disorder (**TUD**; OR_F_ = 1.17 [1.13, 2.21])), musculoskeletal conditions (e.g., osteoarthritis (OR_F_ = 1.07 [1.03, 1.11])), dermatologic conditions (e.g., degenerative skin conditions (OR_F_ = 1.12 [1.06, 1.18])), genitourinary conditions (e.g., end stage renal disease (OR_F_ = 1.16 [1.08, 1.24])), and circulatory conditions (e.g., heart failure (OR_F_ = 1.08 [1.04, 1.11]); **Figure 4B**). Four of these associations were not significant in the sex-combined PGS analyses (**Table S7**).

In females, we also observed 40 (74%) negative associations (i.e., linked to reduced risk for medical conditions), particularly for *drinking frequency* (n = 9)*, drinking quantity* (n = 14) and *AUDIT total* (n = 14) PGS (**Figure 4A**). These included associations with lower risk for digestive diseases (e.g., chronic liver disease and cirrhosis (OR_F_ = 0.91 [0.85, 0.97])), neurological (e.g., sleep apnea (OR_F_ = 0.88 [0.84, 0.92])), endocrine (e.g., type 2 diabetes (OR_F_ = 0.87 [0.83, 0.91])), circulatory conditions (e.g., ischemic heart disease (OR_F_ = 0.92 [0.88, 0.96])), dermatologic conditions (e.g., psoriatic arthropathy (OR_F_ = 0.77 [0.61, 0.93])), genitourinary (e.g., renal failure (OR_F_ = 0.95 [0.93, 0.97])), and hematopoietic conditions (e.g., diseases of white blood cells (OR_F_ = 0.92 [0.86, 0.98]); **Figure 4B**, **Table S7**). Eleven of these associations were not significant in the sex-combined PGS analyses (**Table S7**), reflecting the importance of conducting sex-specific analyses.

In the male PGS analyses, 31 (81.6%) of the sex-specific associations were positive, particularly for *beer quantity* (n = 19) and *PAU* (n = 6) PGSs (**Figure 4A**). These included associations with higher risk for SUDs (e.g., TUD (OR_M_ = 1.21 [1.15, 1.27])), mental conditions (e.g., anxiety (OR_M_ = 1.12 [1.08, 1.16]), as well as medical comorbidities that result from substance use, such as digestive conditions (e.g., alcoholic liver damage (OR_M_ = 1.59 [1.43, 1.75])), genitourinary conditions (e.g., renal failure (OR_M_ = 1.09 [1.05, 1.13])), hematopoietic conditions (e.g., thrombocytopenia (OR_M_ = 1.16 [1.08, 1.24])), infectious diseases (e.g., viral hepatitis C (OR_M_ = 1.37 [1.25, 1.49])), neoplasm diseases (e.g., liver cancer (OR_M_ = 1.31 [1.15, 1.47])), and respiratory diseases (e.g., chronic airway obstruction (OR_M_ = 1.14 [1.08, 1.2]); **Figure 4B**, **Table S7**). Four of these associations were not significant in the sex-combined PGS analyses (**Table S7**).

In males, only seven (18.4%) sex-specific associations were negative, including associations with lower risk for digestive diseases (i.e., celiac disease (OR_M_ = 0.66 [0.5, 0.82])) and endocrine and metabolic conditions (i.e., type 1 diabetes (OR_M_ = 0.91 [0.85, 0.97])), and neoplasm diseases (i.e., hemangioma and lymphangioma (OR_M_ = 0.77 [0.63, 0.91])). Two of these associations were not significant in the sex-combined PGS analyses (**Table S7**).

Finally, five associations were positively associated in both sexes but showed significantly stronger associations in males, particularly for *PAU* (n = 4) and *beer quantity* (n = 1) PGS and substance use traits (e.g., AUD (OR_F_ = 1.18 [1.08, 1.28] vs. OR_M_ = 1.41 [1.35, 1.47]); **Figure 4A-B**, **Table S7**).

To assess whether sex-specific PGS improved prediction of AUD diagnosis, we compared the variance explained (Nagelkerke *R²*) by sex-specific and sex-combined PGS. Consistent with our third hypothesis, sex-specific PGSs did not outperform the sex-combined PGSs in predicting AUD in either the female or male samples (**Figure 5**, **Tables S7-8**). In fact, several sex-specific PGSs (i.e., *drinking with meals, decreased drinking, spirits, red* and *white wine quantity and AUDIT total*) were not significantly associated with AUD in either sex (**Table S7**).

**Figure 5.**
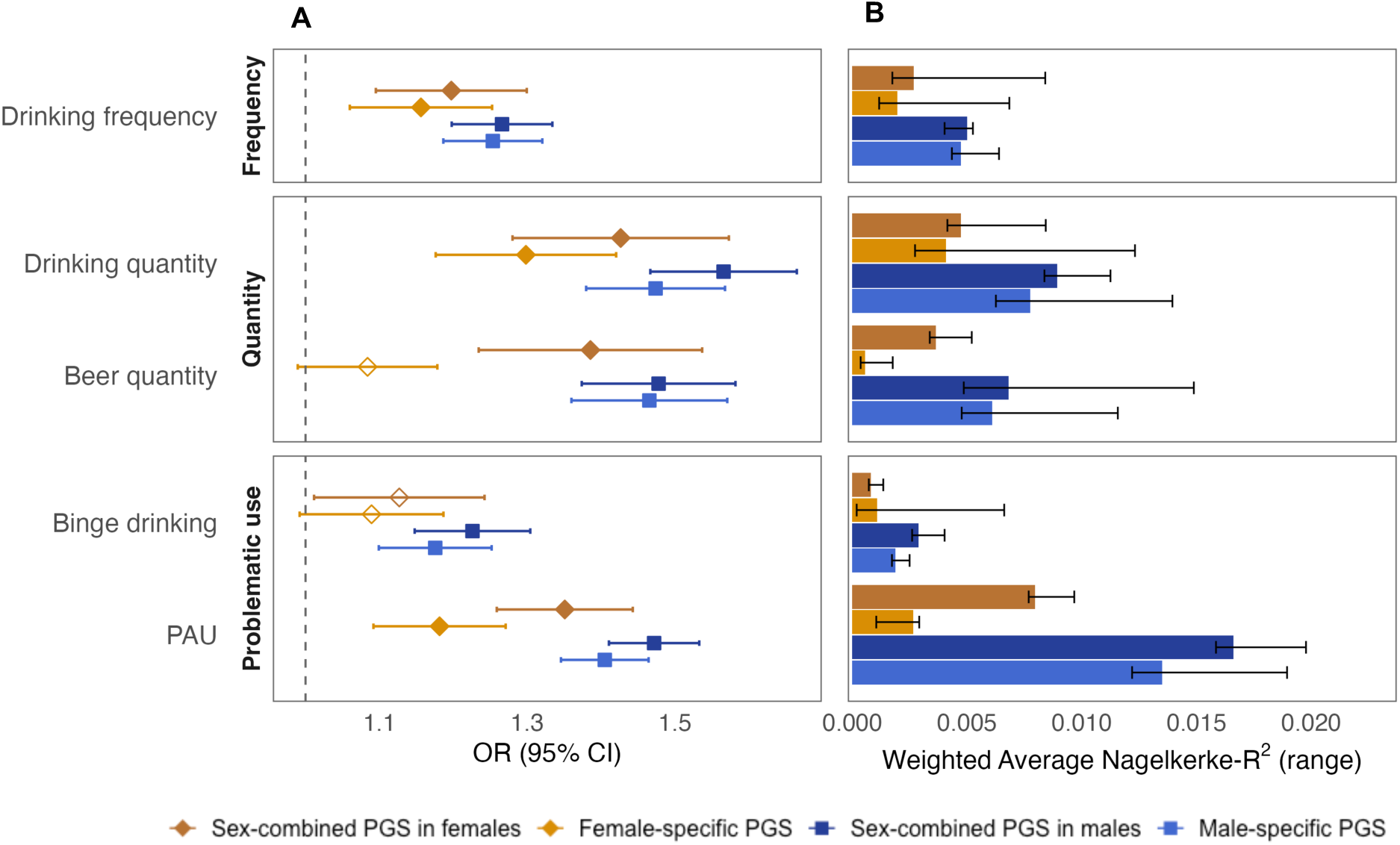
Predictive ability assessment of sex-specific and sex-combined polygenic scores (PGS) for alcohol use phenotypes on alcohol use disorder (AUD) diagnosis in BioVU and PMBB. **A.** Odds ratios (OR) and 95% confidence intervals for the association between each alcohol use phenotype PGS and AUD Phecode in sex-stratified target samples. **B.** Weighted average Nagelkerke-R² values for each PGS in predicting AUD in BioVU and PMBB. The weighted average Nagelkerke *R²* across sites was calculated with the formula Nagelkerke-*R²_WA_* = ∑ w_i_\**R*_i_*²*/∑ w_i_. The range corresponds to minimum and maximum Nagelkerke *R²* across sites (see **Table S8**). Male-specific PGS are in blue squares, sex-combined PGS in males are in dark blue squares, female-specific PGS are in amber triangles, and sex-combined PGS in females are in dark amber triangles. Only alcohol use phenotype for which the PGS showed a significant association with AUD in either of the sex-specific PGSs are displayed. *Drinking with meals, decreased drinking*, *spirits, red* and *white wine quantity* and *AUDIT total* PGSs were not significantly associated with AUD in either sex (**Table S7**).

## DISCUSSION

Sex differences in alcohol use patterns and trajectories are well documented, but the biological mechanisms underpinning these differences are relatively unknown^7^. Using sex-stratified GWAS of alcohol use behaviors spanning measures of frequency, quantity and problematic use, we characterized their genetic architecture separately in females and males. We evaluated both *quantitative* sex differences (i.e., the magnitude of genetic influence by sex) and *qualitative* differences (i.e., specific genetic variants). Our findings supported our original hypotheses: (i) quantitative sex differences were broadly more pronounced for drinking quantity phenotypes, but less notable for frequency or problematic use, (ii) externalizing comorbidities showed stronger genetic correlations in males with less evidence for stronger genetic correlations with internalizing comorbidities in females, and (iii) sex-specific PGSs revealed associations with sex-specific comorbidities, some of which were masked in sex-agnostic analyses. We argue that phenotypes strongly influenced by cultural and social differences, particularly the choice of beverage, shapes these sex-specific comorbidity patterns, and outline considerations for future sex-aware studies of alcohol use phenotypes.

Genetic and environmental factors jointly shape alcohol use behaviors. Twin studies have repeatedly shown that alcohol use behaviors are heritable, but evidence for large quantitative sex differences in heritability remains limited (40-45% in females, 50-52% in males;^21,54–58^. Our GWAS results are largely consistent with these findings. We identified significant *h^2^_SNP_* for all 11 phenotypes, with similar estimates across sexes for most traits. This aligns with prior UK Biobank analyses showing minimal cross-sex divergence in *h^2^_SNP_* estimates across hundreds of other complex traits, including psychiatric and behavioral phenotypes^59^. Thus, data both from family studies and GWAS suggest that the overall genetic contribution to alcohol use behaviors is broadly similar by sex.

Despite this broad similarity, important sex differences still emerged in our study for *beer quantity* and *PAU*, which showed significantly higher *h^2^_SNP_* in males compared to females. Because heritability depends on the study population and measurement^59,60^, these gaps may reflect non-genetic influences, such as sample imbalance (e.g., 496,548 males vs. 143,198 females for *PAU*), differences in endorsement likely reflecting differences in social norms (e.g., UK Biobank male participants reported drinking ∼5x more beer compared to females), or reporting biases (e.g., due to stigma surrounding alcohol use in females)^61,62^. Therefore, these differences should be interpreted with caution until replicated in balanced cohorts with harmonized measures.

Over the past decade, sex-agnostic GWASs have identified hundreds of loci associated with aspects of alcohol use behaviors^40,63–65^. When conducting GWAS in sex-stratified cohorts, we identified that sex-differentiated effects existed but were sparse. While the magnitude of effect across variants was largely concordant across sexes, we found four loci with significant sex differences: a female-specific association in *IZUMO1* (rs838147) for *drinking frequency* and *quantity*, and three male-specific associations in *ADH1B* (rs1229984), *KLB* (rs28712821), and *FTO* (rs1421085) for *beer quantity* and/or *PAU*. However, there are two caveats to note. First, it is possible that differences in ascertainment biases may be again conflating the findings. For example, *ADH1B*, one of the most well-established genes implicated in alcohol use and AUD, shows robust associations with several alcohol use phenotypes in females, suggesting that the male-specific association observed for *beer quantity* may instead reflect reduced power in the female sample for this particular phenotype rather than true sex-differentiated genetic effects. In addition, a recent study using UK Biobank data identified a BMI-increasing *FTO* allele (rs10468280) to be at a higher frequency in males and more strongly associated with BMI in males, consistent with sex-differential participation bias^66^. Similarly, we identified an *FTO* variant in high LD (r^2^ = 0.9) with rs10468280 that showed sex-differentiated genetic effects in males for *PAU*, which may reflect similar selection biases rather than biology. Second, sex-stratified GWASs can only substitute genotype-by-sex interaction tests when covariate interactions and variance differences are equivalent in the two sexes. Therefore, individual-level data and genotype-by-sex interaction test will be ultimately needed to evaluate whether these reported sex differences are driven by genetic or non-genetic factors^67^. Overall, the absence of sex-differentiated loci for the remaining alcohol phenotypes may partly reflect limited statistical power rather than proof of homogeneity^68^. Indeed, sex-differentiated effects are now beginning to emerge for large-scale GWAS of somatic traits^69^. It is plausible that, for alcohol use phenotypes, as seen for other complex behavioral and psychiatric traits^70^, much larger and more balanced samples will be needed to detect sex-differentiated genetic effects, which, even if small, could be informative for mechanism and comorbidity.

Equal *h^2^_SNP_* across sexes and limited genome-wide sex-differentiated effects do not imply that the underlying polygenic architecture is shared in females and males^41^. In our data, between-sex genetic correlations significantly departed from 1.0, suggesting that sex-differentiated genetic effects for the alcohol phenotypes tested are prevalent but small in magnitude and distributed across the genome^22,26,71^. Mean *r_g_* values were generally lower for quantity phenotypes (mean *r_g_* = 0.87 ± 0.06 [frequency], 0.78 ± 0.06 [quantity] and 0.85 ± 0.08 [problematic use]). Differences were particularly pronounced for certain quantity measures, particularly *spirit quantity and beer quantity* (*r_g_* = 0.68 and 0.70), but less for others, such as *red wine quantity* (*r_g_* = 0.89), perhaps reflecting cultural or social differences in drinking behaviors around these specific beverages.

Cross-trait genetic correlations among alcohol phenotypes also differed between sexes: 43% of pairwise comparisons showed sex divergence, with some showing opposite patterns in males and in females. For instance, *drinking quantity* and *drinking with meals* were negatively genetically correlated in males, but positively in females. Males reported consuming larger quantities of alcohol on average (2.81 ± 1.00 vs. 2.12 ± 1.01), yet they were less likely to drink with meals (58.5% vs. 77.6%) compared to females. Thus, in males, drinking quantity likely reflects higher-volume drinking episodes outside mealtimes (often involving beer). Furthermore, even with the same number of drinks, females can reach higher blood alcohol concentrations due to lower total body water content^17,27^. These patterns in behavior, physiological and reporting differences can bias genetic correlation estimates. Scaling measures of intake (g of ethanol/kg), modeling beverage and meal context, and testing methods that use multimodal techniques (e.g., Genomic SEM) to account for participation bias by sex^26,66^ could clarify whether the observed sex-dependent patterns reflect biological or environmental differences.

When examining the relationship between female and male alcohol use behaviors and other comorbid traits, 18.6% of the genetic correlations that were significant in either males or females showed significant sex differences. We identified patterns partially consistent with our second hypothesis. In males, genetic correlations with externalizing traits (e.g., SUDs, ADHD, externalizing, extraversion or delay discounting) were stronger for *drinking quantity, beer quantity* and *PAU* compared to females. In females, we identified a single stronger genetic correlation with an internalizing trait (i.e., self-harm) for *beer quantity*, suggesting that the previously observed higher comorbidity with internalizing psychopathology in females^28,72^ may be largely influenced by environmental or social factors rather than genetic influences. These patterns are consistent with epidemiologic and twin evidence^73,74^, reinforcing that AUD likely develops through partly different etiologic pathways in females and males.

Once more, the largest sex differences were found for overall drinking quantity rather than frequency or problematic use phenotypes and varied by beverage type. For example, *beer quantity* showed *positive* genetic correlations for externalizing traits in males, whereas *spirit quantity* showed *positive* genetic correlations for the same externalizing traits in females. In contrast, *red* and *white wine quantity* in males showed *negative* genetic correlations with externalizing traits. These findings may reflect cultural factors, including social norms, surrounding alcohol consumption, rather than purely biological differences. Furthermore, we identified positive genetic correlations with *better* socioeconomic outcomes for overall drinking quantity in females only, and for red and white wine consumption in both sexes. This suggests that the counterintuitive positive genetic correlations that we and others have previously noted between *better* socio-educational outcomes and alcohol consumption in sex-combined samples, which likely reflects collider bias from nonrandom sampling in biobank studies^75–78^, may be largely driven by females and/or by specific beverage types, particularly wine, which is often a marker for higher socioeconomic status. Future work should account for beverage preference to avoid masking or mischaracterizing sex-differentiated effects.

In hospital-based cohorts, PGS analyses revealed sex-specific associations with alcohol-related medical comorbidities. Across BioVU and PMBB, 19.9% PGS-trait comparisons showed significant sex differences. These differences are unlikely to be simply a power artifact, because target-sample sizes by sex were comparable for most traits. In females, female-specific PGS were associated with increased risk of osteoarthritis, degenerative skin conditions, cardiomegaly, and end-stage renal disease, among others. In contrast, male-specific PGS associations concentrated in SUDs, alcohol-related medical sequelae (e.g., alcoholic liver damage, liver cancer), and other externalizing-related traits (i.e., fracture of lower limb), consistent with the epidemiological literature reporting heavier drinking and higher externalizing behaviors in males^79,80^. Strikingly, some of these associations, such as the female-specific association with osteoarthritis and the male-specific association with fracture of the lower limb, were not detected in sex-combined PGS analyses, highlighting the importance of sex-stratified approaches for uncovering sex-specific comorbidity profiles of alcohol use behaviors that were masked in sex-combined analyses. We also observed *negative* PGS associations, primary between *drinking frequency* PGS and some medical conditions, including circulatory, metabolic, respiratory and digestive conditions (**Figure S7**), which persisted after covarying for AUD diagnosis in sensitivity analyses (**Table S9**). This pattern of paradoxical associations is unlikely to reflect true protective biological effects. In fact, phenotypic PheWAS of AUD in these same clinical populations did not recapitulate these patterns, instead showing positive associations with these conditions (**Table S10**). Together, these findings suggest that the apparent pattern of *protective* associations may reflect participation biases from the healthier than average UK Biobank^81^ discovery GWAS used to construct the PGS, rather than true inverse biological relationships.

Sex-stratified PGS did not outperform sex-combined PGS for AUD prediction, consistent with the idea that pooling females and males to compute PGSs often yields more precise SNP weights when cross-sex genetic overlap is substantial (*r_g_* 0.7–0.9)^82^. As larger and more balanced sex-stratified GWAS become available, emerging approaches that jointly model genetic effects across sexes (e.g., PRS-CSx)^74,82,83^ can be applied to examine whether these approaches show better predictive performance in females and males.

Our study is not without limitations. First, GWAS are disproportionately male-dominant, especially for PAU, reflecting historical prevalence differences or ascertainment bias. This sample imbalance can result in power differences and inflate apparent male effects. Second, many sex-stratified alcohol GWAS were derived from volunteer cohorts (e.g., UK Biobank), where sex-differential participation can induce or magnify cross-trait correlations^60,84^. Consumption phenotypes from volunteer cohorts may benefit from multivariate frameworks (e.g., Genomic SEM) to partial out shared structure not directly related to AUD liability, as we have shown in prior work^25^. Third, sex differences are unlikely to be explained solely by common autosomal variation. Although GWAS using UKB Biobank and 23andMe data included the X chromosome, X-aware tools (e.g., methods accounting for X-inactivation, such as GXwasR^85^, and rare-variant models are needed to test whether non-autosomal or rare variation contributes to sex differences^60^. Finally, gene-by-environment interactions likely contribute to sex differences: genetic influences on alcohol use often strengthen in permissive contexts (e.g., peer drinking, low relationship/support), and attenuate in restrictive contexts (e.g., non-drinking peers or medical advice)^86^, with some evidence showing that this effect is stronger in females^20,87^. In addition, social norms and behaviors surrounding alcohol consumption in the UK, where much of our data originate, likely differ across countries, ages and historical moments. Future work should include frameworks for quantifying and estimating polygenic GxE by sex, as well as incorporate data from diverse sociocultural settings^88^.

In summary, sex differences in the genetic architecture of alcohol use phenotypes are polygenic and subtle, with little evidence for single variants with large sex-differentiated effects. Patterns of genetic correlation with psychiatric traits and medical outcomes were sex-specific, even when SNP-based heritability and genetic loci were mostly similar across sexes. The most pronounced sex differences were observed for drinking quantity measures and were influenced by beverage type. This work continues to motivate sex-aware analyses that consider the spectrum of alcohol use behaviors and environmental information in larger, balanced cohorts. Adopting these practices will enable future genetic studies that address, rather than ignore, AUD heterogeneity in females and males.

## Data Availability

All data produced in the present work are contained in the manuscript

## Acknowledgements

This work was supported by National Institutes of Health grant numbers DP1DA054394 (JJM, SSR), T32GM139790 (JG), R25MH081482 (LV-R) and K01DA051759 (ECJ), by the National Institute of Mental Health grant numbers R01MH137219-01 (SSR), R01MH137220-01 (LKD) and R01-MH137212-01 (RLK), by the Tobacco-Related Disease Research Program grant numbers T35FT9589 (LV-R), T33KT6694 (NC-K) and T35MS9695 (NC-K), and the National Institute on Alcohol Abuse and Alcoholism grant number L40AA031140 (NC-K).

We would like to thank the research participants and employees of 23andMe Research Institute for making this work possible. The following members of the 23andMe Research Team contributed to this study: Stella Aslibekyan, Adam Auton, Elizabeth Babalola, Robert K. Bell, Jessica Bielenberg, Ninad S. Chaudhary, Zayn Cochinwala, Sayantan Das, Emily DelloRusso, Payam Dibaeinia, Sarah L. Elson, Nicholas Eriksson, Chris Eijsbouts, Teresa Filshtein, Pierre Fontanillas, Davide Foletti, Will Freyman, Zach Fuller, Julie M. Granka, Chris German, Éadaoin Harney, Alejandro Hernandez, Barry Hicks, David A. Hinds, M. Reza Jabalameli, Ethan M. Jewett, Yunxuan Jiang, Sotiris Karagounis, Lucy Kaufmann, Matt Kmiecik, Katelyn Kukar, Alan Kwong, Keng-Han Lin, Yanyu Liang, Bianca A. Llamas, Aly Khan, Steven J. Micheletti, Matthew H. McIntyre, Meghan E. Moreno, Priyanka Nandakumar, Dominique T. Nguyen, Jared O’Connell, Steve Pitts, G. David Poznik, Alexandra Reynoso, Shubham Saini, Morgan Schumacher, Leah Selcer, Anjali J. Shastri, Jingchunzi Shi, Suyash Shringarpure, Keaton Stagaman, Teague Sterling, Qiaojuan Jane Su, Joyce Y. Tung, Susana A. Tat, Vinh Tran, Xin Wang, Wei Wang, Catherine H. Weldon, Amy L. Williams, Peter Wilton.

We acknowledge the Penn Medicine BioBank (PMBB) for providing data and thank the patient- participants of Penn Medicine who consented to participate in this research program. We would also like to thank the Penn Medicine BioBank team and Regeneron Genetics Center for providing genetic variant data for analysis. The PMBB is approved under IRB protocol# 813913 and supported by Perelman School of Medicine at University of Pennsylvania, a gift from the Smilow family, and the National Center for Advancing Translational Sciences of the National Institutes of Health under CTSA award number UL1TR001878.

## Conflict of interest

S.L.E. and P.F. were employed by and held stock or stock options in 23andMe, Inc. at the time of the study. N.C.K. is consultant and holds stock options in CARI Health, Inc. All other authors report no biomedical financial interests or potential conflicts of interest.

## Notes

### Summary of Updates

The funding source test was extended to include missing grants related to this work.

