## Supplementary Figures for "Sex-Specific Genetic Architecture and Comorbidities of Alcohol Use Behaviors"

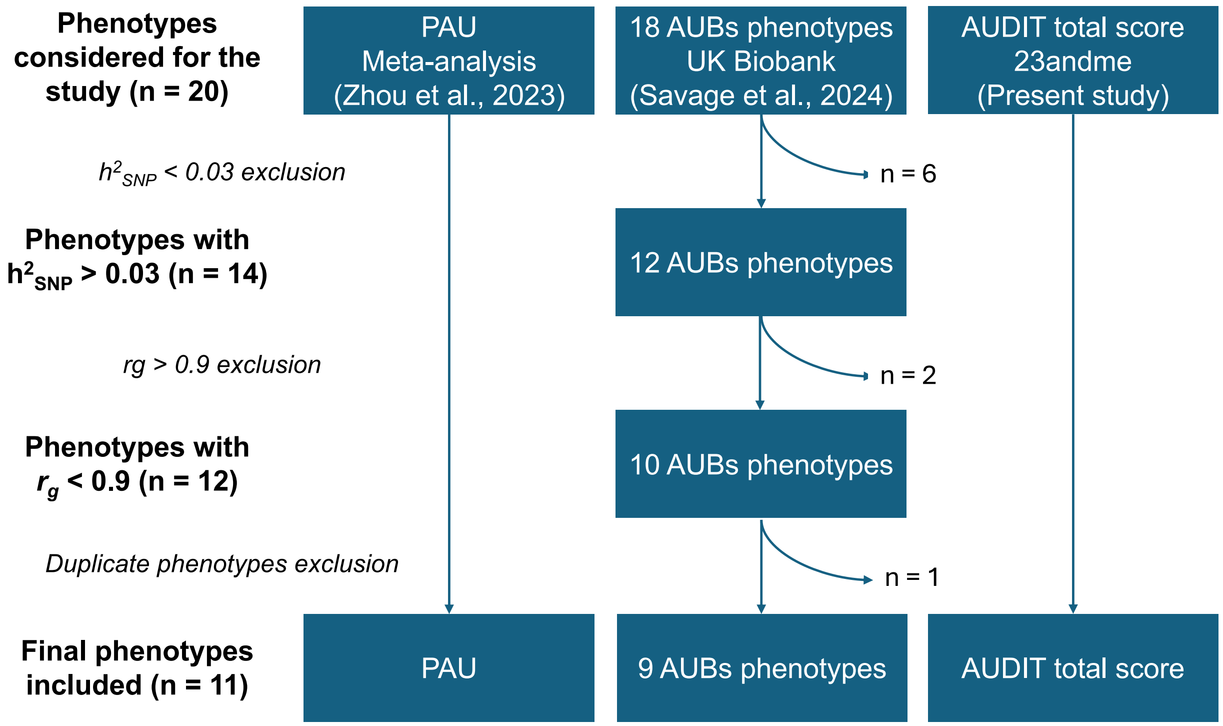

**Figure S1.** Flowchart overview of data selection for the study. PAU: Problematic alcohol use. AUBs: Alcohol use behaviors. Phenotypes excluded for SNP-based heritability (***$h^2_{SNP}$***) < 0.03: *abuse* (Harmful/risky use), *advice* (received counseling from medical practitioner about alcohol use), *anyclin* (any clinically significant event related to alcohol misuse), *broad_aud* (broad alcohol use disorder definition), *increasedrink* (drinking has increased over the past 10 years), *quantfwine* (quantity of fortified wine consumed per month). Phenotypes excluded for $r_g$ > 0.9: *auditc_ln* (AUDIT-C score), *pershistory* (self-report of ever having physical addiction to alcohol). AUDIT-P was included in PAU; therefore, we excluded *mh_auditp* (AUDIT-P score) from the analyses.

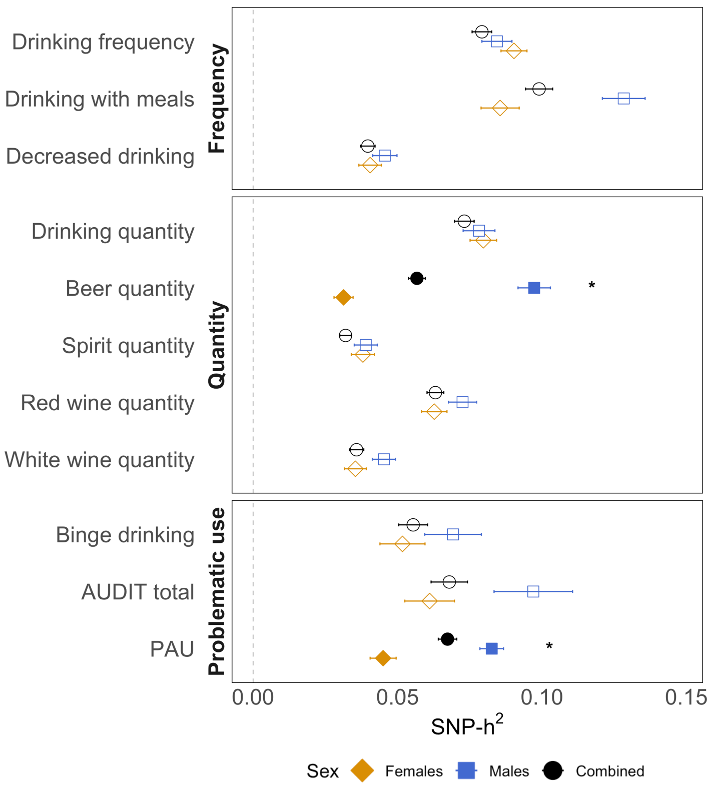

**Figure S2.** Overview of $h^2_{SNP}$ estimates (observed scale) across the 11 alcohol use phenotypes. Sex-combined results are shown in black dots, males are represented in blue squares, and females are represented in yellow triangles. Alcohol use phenotypes are grouped by category: frequency, quantity, and problematic use. Asterisks and filled symbols indicate significant sex differences in $h^2_{SNP}$ estimates (FDR corrected $p < 0.05$).

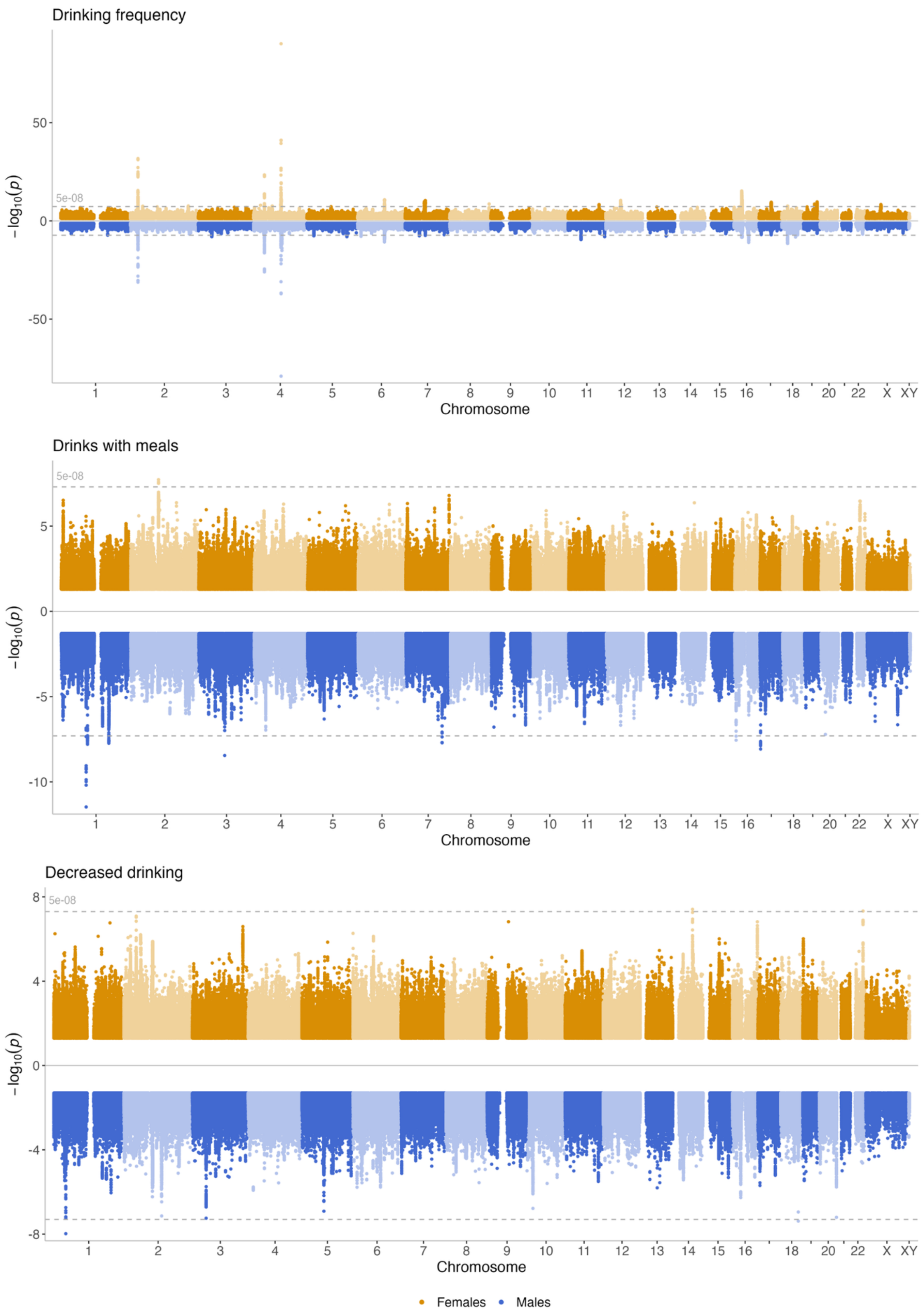
Drinking frequency
Drinks with meals
Decreased drinking
5e-08
$-\log_{10}(p)$
Chromosome
Females
Males

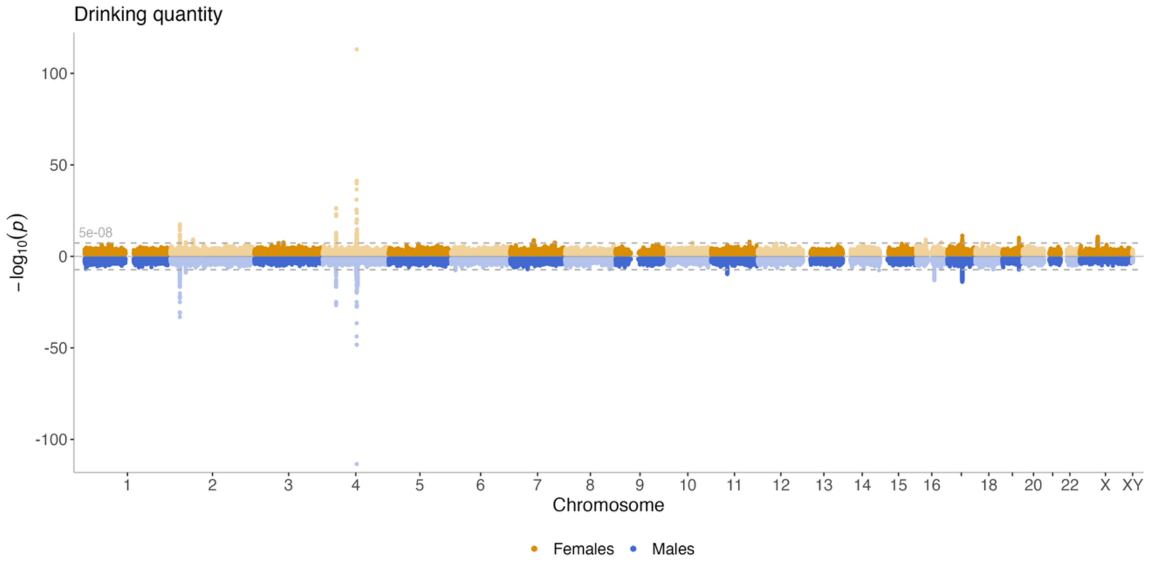
Drinking quantity
100
50
0
-50
-100
5e-08
$-\log_{10}(p)$
1 2 3 4 5 6 7 8 9 10 11 12 13 14 15 16 18 20 22 X XY
Chromosome
Females
Males

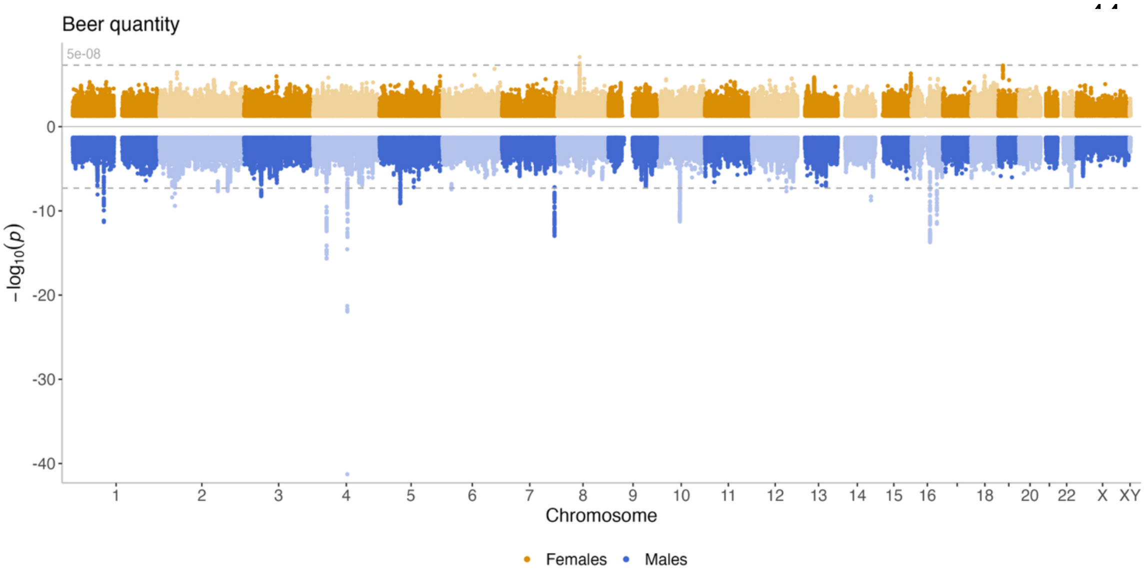
Beer quantity
5e-08
0
-10
-20
-30
-40
$-\log_{10}(p)$
1 2 3 4 5 6 7 8 9 10 11 12 13 14 15 16 18 20 22 X XY
Chromosome
Females
Males

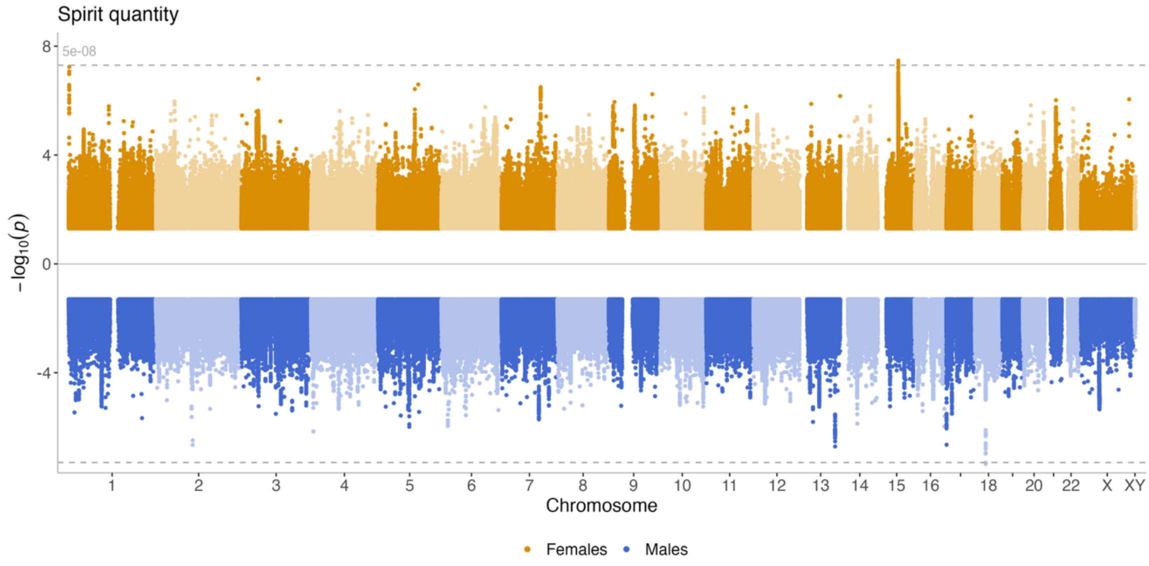
Spirit quantity
8
4
0
-4
5e-08
$-\log_{10}(p)$
1 2 3 4 5 6 7 8 9 10 11 12 13 14 15 16 18 20 22 X XY
Chromosome
Females
Males

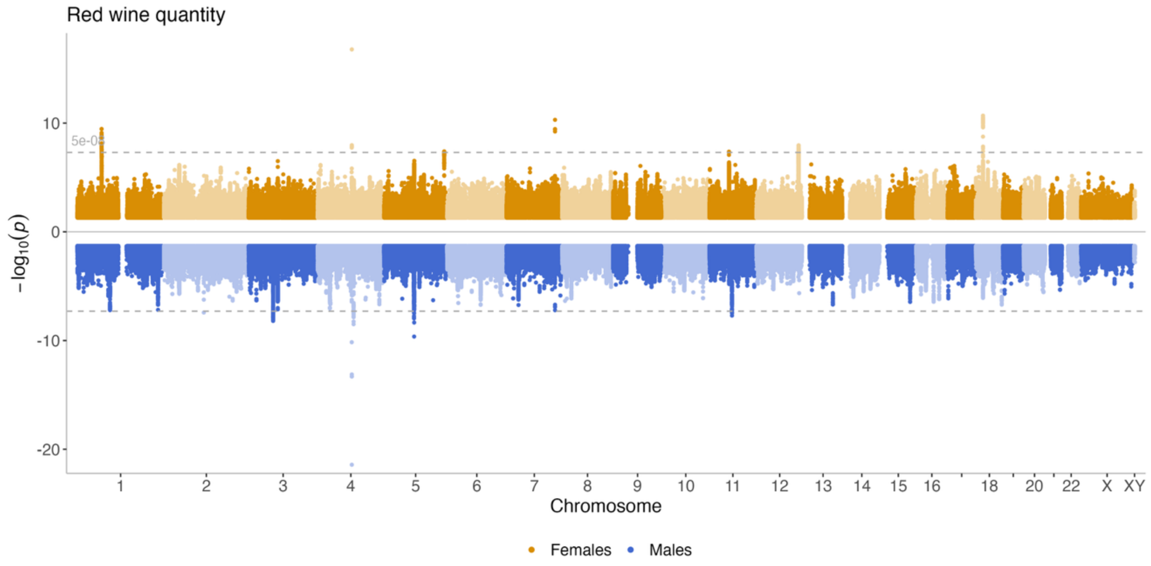
Red wine quantity
5e-08
10
0
-10
-20
−log10(p)
18
20
22
X
XY
Chromosome
Females
Males

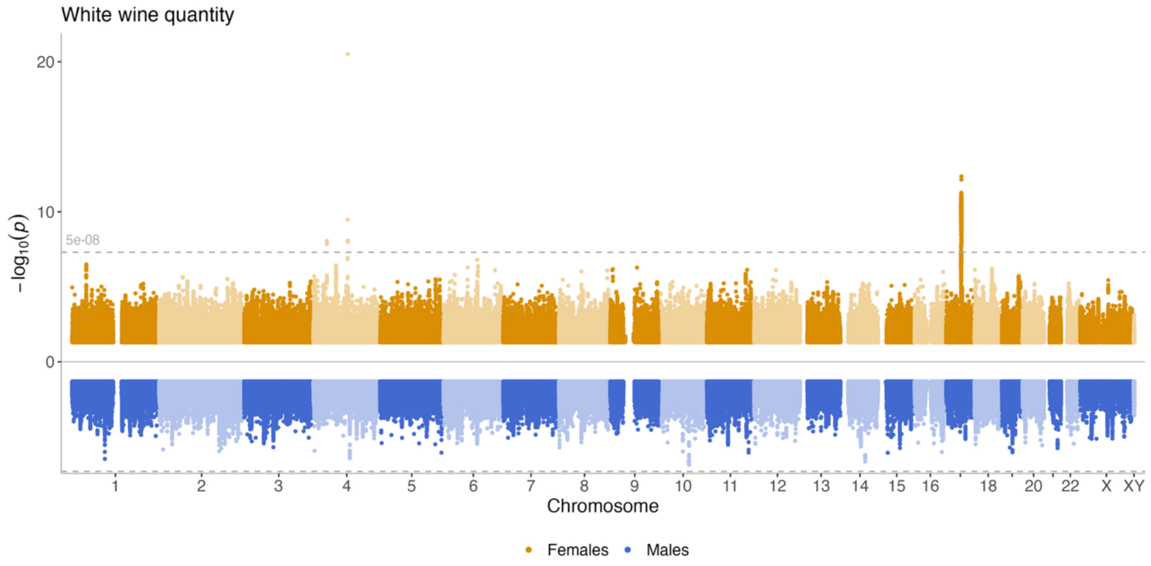
White wine quantity
20
10
5e-08
0
−log10(p)
18
20
22
X
XY
Chromosome
Females
Males

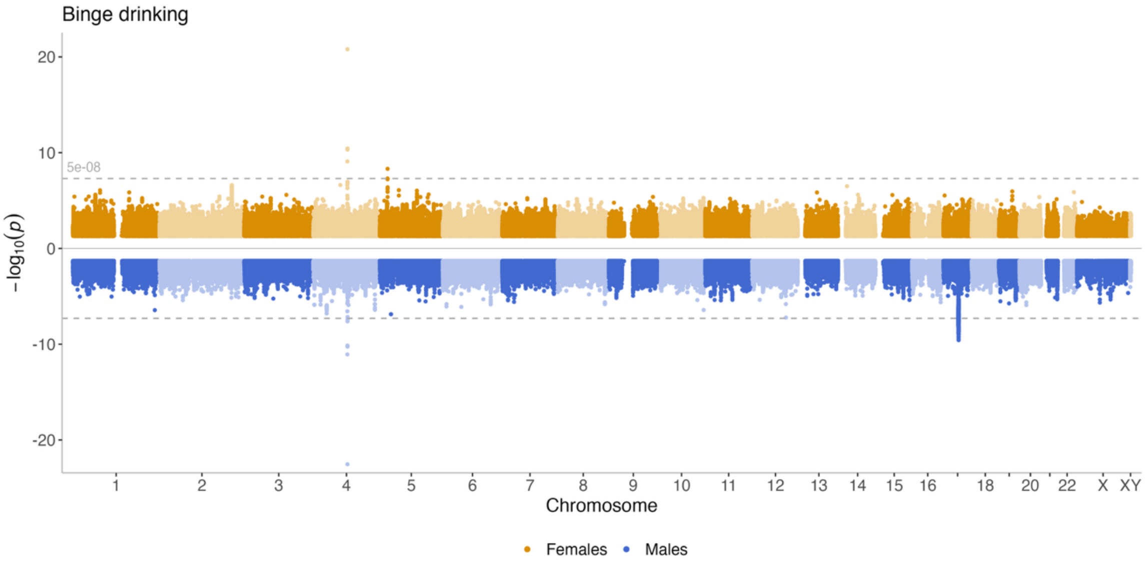
Binge drinking
20
10
5e-08
0
-10
-20
−log10(p)
18
20
22
X
XY
Chromosome
Females
Males

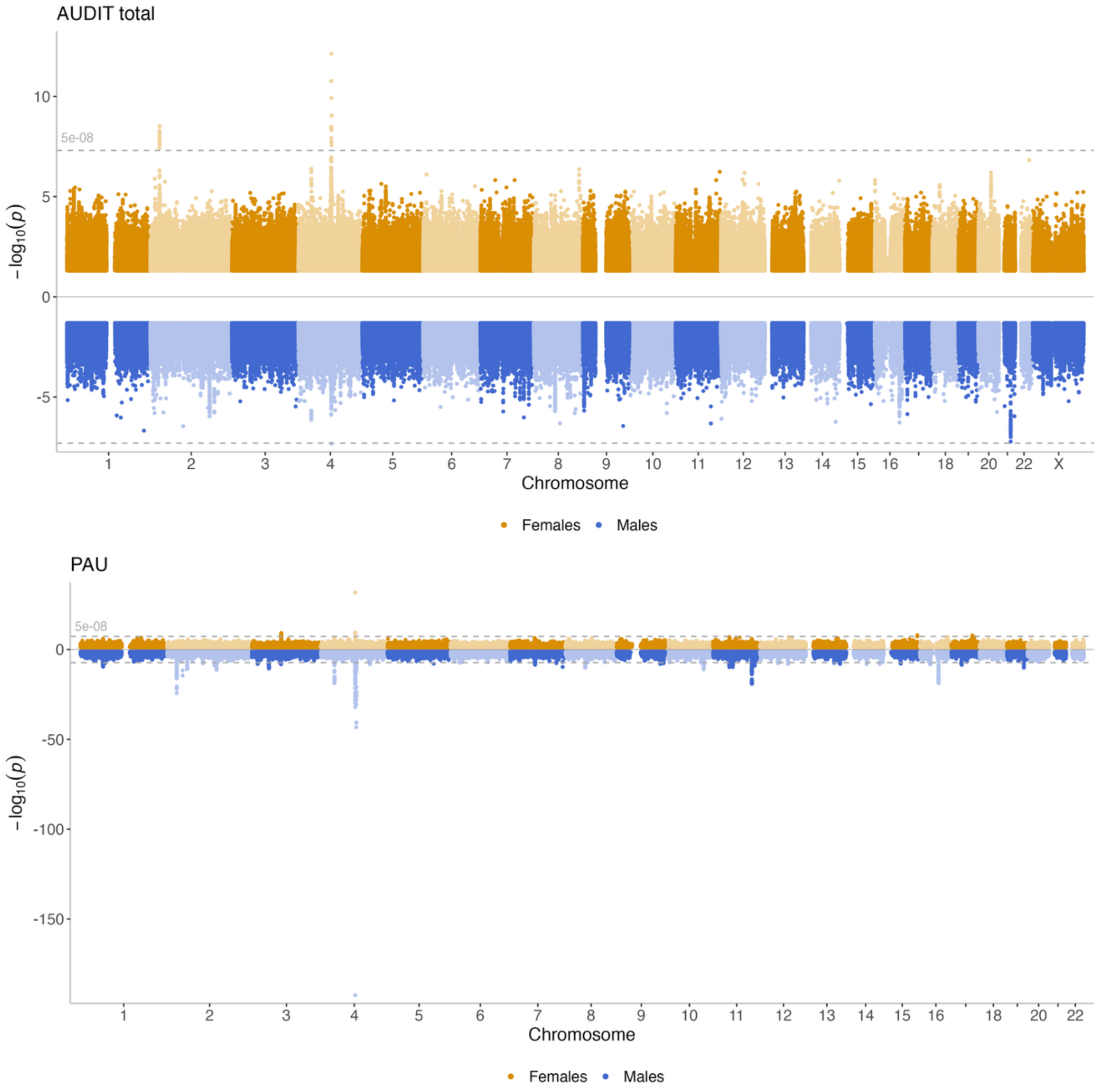

**Figure S3.** Miami plots for the 11 alcohol use phenotypes included in the study. The -$\log_{10}(p)$ for the female GWASs are plotted in the upper panel in yellow, and the -$\log_{10}(p)$ for the male GWASs are plotted in the lower panel in blue.

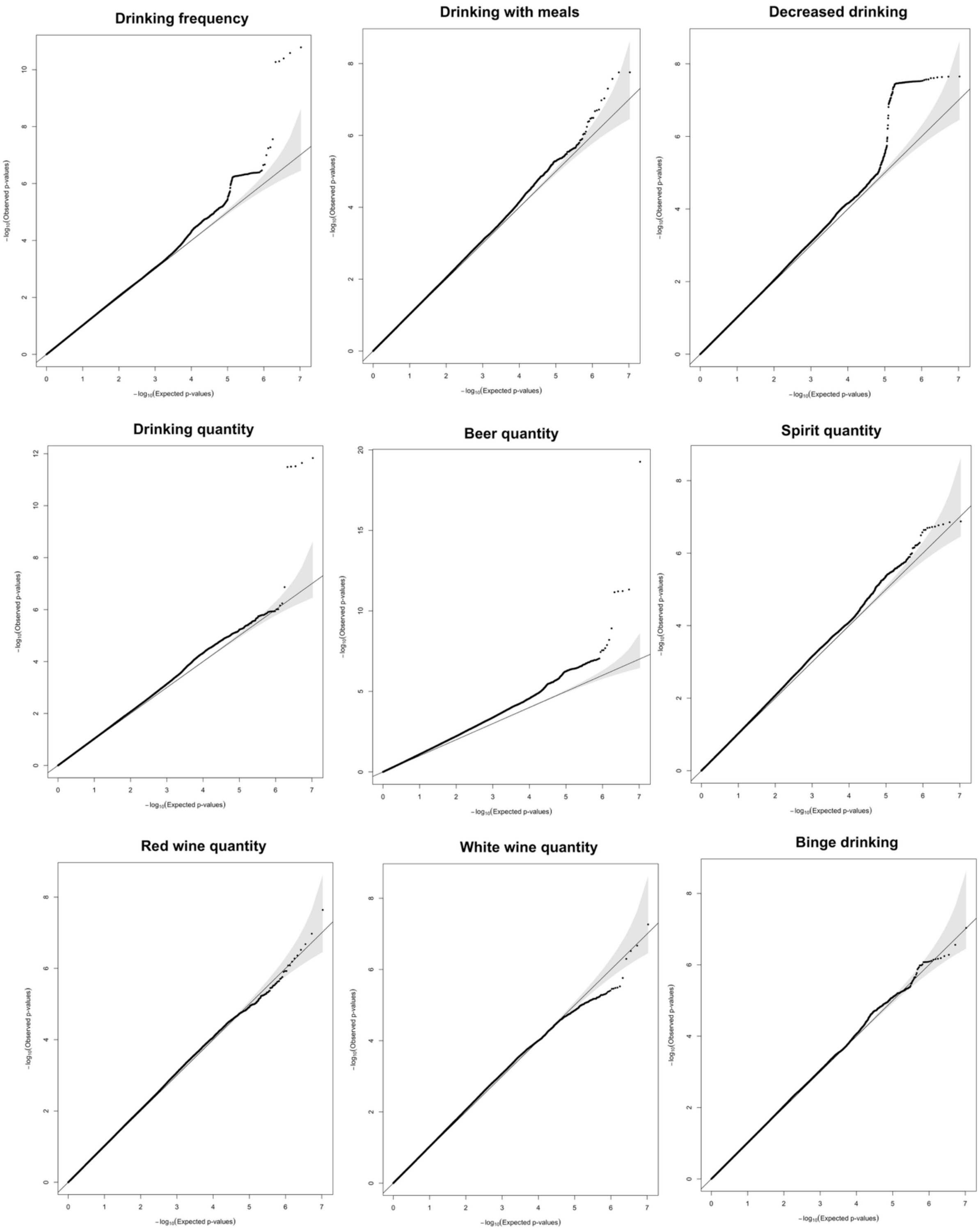
Drinking frequency
Drinking with meals
Decreased drinking
Drinking quantity
Beer quantity
Spirit quantity
Red wine quantity
White wine quantity
Binge drinking
$-\log_{10}$(Observed p-values)
$-\log_{10}$(Expected p-values)

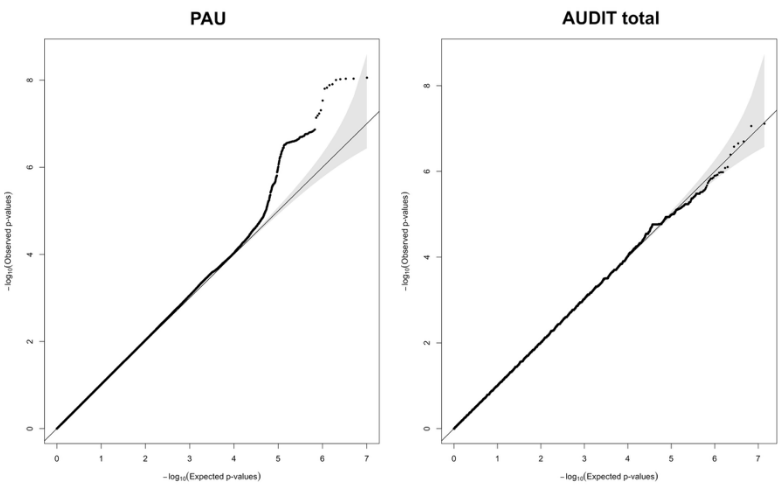

**Figure S4.** We derived quantile-quantile (**QQ**) plots of the -log$_{10}$($p$) distribution to inspect and visualize differences in SNP effect sizes between females and males. Theoretical -log$_{10}$($p$) are shown on the x-axis, and experimental -log$_{10}$($p$) are shown on the y-axis. The X:Y line is plotted as a black dashed line, and the grey area represents the 95% confidence interval.

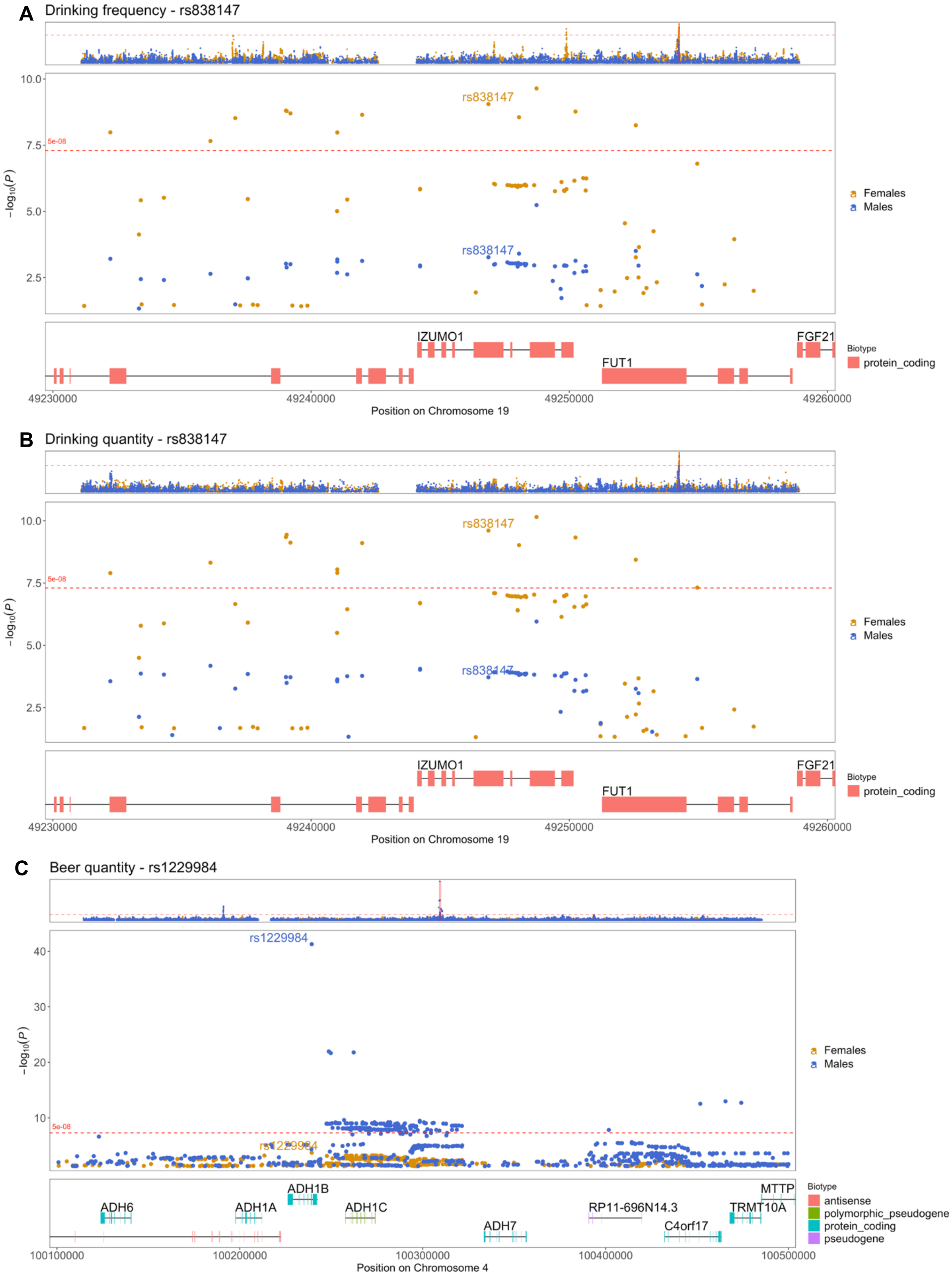
A
Drinking frequency - rs838147
rs838147
rs838147
5e-08
Females
Males
IZUMO1
FGF21
FUT1
Biotype
protein_coding
Position on Chromosome 19
B
Drinking quantity - rs838147
rs838147
rs838147
5e-08
Females
Males
IZUMO1
FGF21
FUT1
Biotype
protein_coding
Position on Chromosome 19
C
Beer quantity - rs1229984
rs1229984
rs1229984
5e-08
Females
Males
ADH1B
ADH6
ADH1A
ADH1C
ADH7
RP11-696N14.3
C4orf17
TRMT10A
MTTP
Biotype
antisense
polymorphic_pseudogene
protein_coding
pseudogene
Position on Chromosome 4

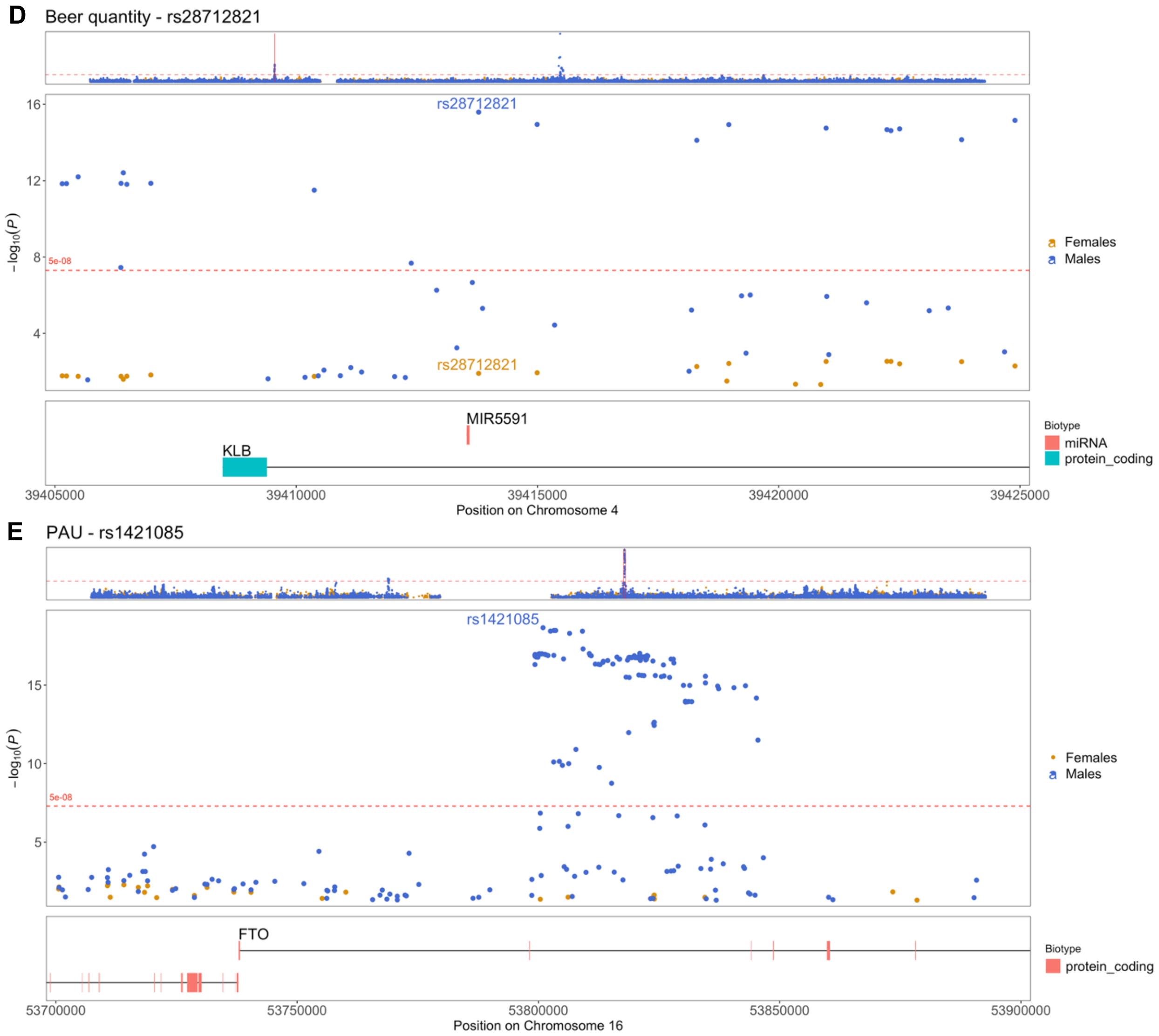

**Figure S5.** Zoom plots for the four independent loci with significant sex-differentiated effects across five alcohol traits. **A-B**. Locus (lead SNP rs838147 in *IZUMO1*) with female-specific effects for both *drinking frequency* (A) and *drinking quantity* (B). **C**. Locus (lead SNPs rs1229984 in *ADH1B*) with male-specific effects for beer quantity. **D**. Locus (lead SNP rs28712821 in *KLB*) with male-specific effects for beer quantity. **E**. Locus (lead SNP rs1421085 in *FTO*) with male-specific effects for PAU.

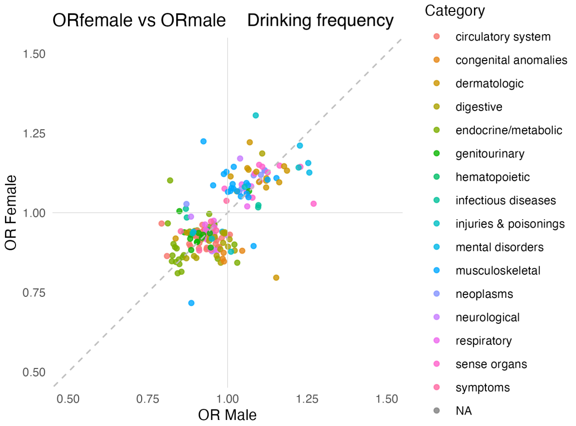
ORfemale vs ORmale Drinking frequency
OR Female
OR Male
0.50
0.75
1.00
1.25
1.50
Category
circulatory system
congenital anomalies
dermatologic
digestive
endocrine/metabolic
genitourinary
hematopoietic
infectious diseases
injuries & poisonings
mental disorders
musculoskeletal
neoplasms
neurological
respiratory
sense organs
symptoms
NA

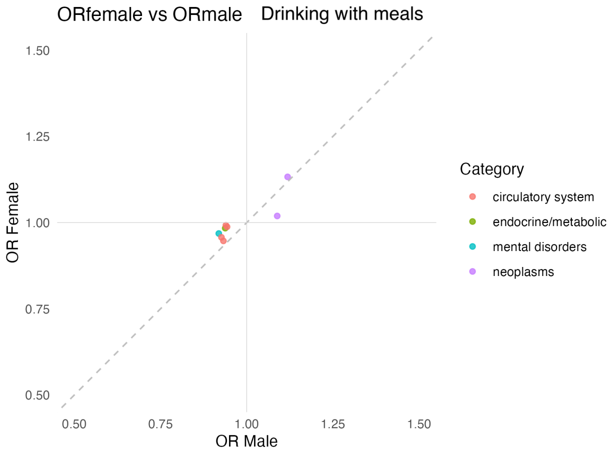
ORfemale vs ORmale Drinking with meals
OR Female
OR Male
0.50
0.75
1.00
1.25
1.50
Category
circulatory system
endocrine/metabolic
mental disorders
neoplasms

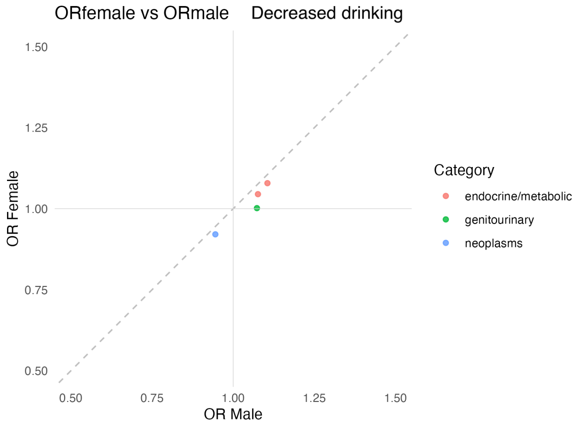
ORfemale vs ORmale Decreased drinking
OR Female
OR Male
0.50
0.75
1.00
1.25
1.50
Category
endocrine/metabolic
genitourinary
neoplasms

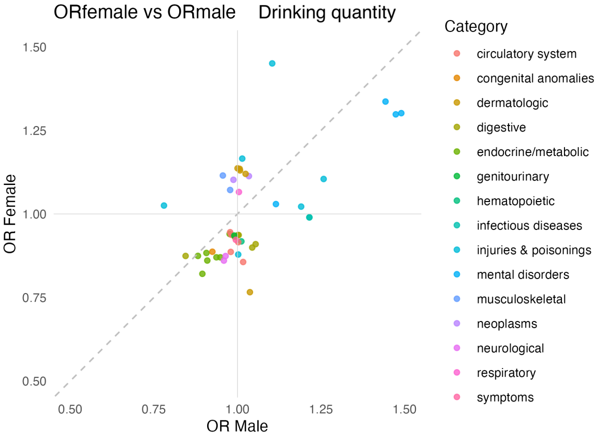
ORfemale vs ORmale Drinking quantity
OR Female
OR Male
0.50
0.75
1.00
1.25
1.50
Category
circulatory system
congenital anomalies
dermatologic
digestive
endocrine/metabolic
genitourinary
hematopoietic
infectious diseases
injuries & poisonings
mental disorders
musculoskeletal
neoplasms
neurological
respiratory
symptoms

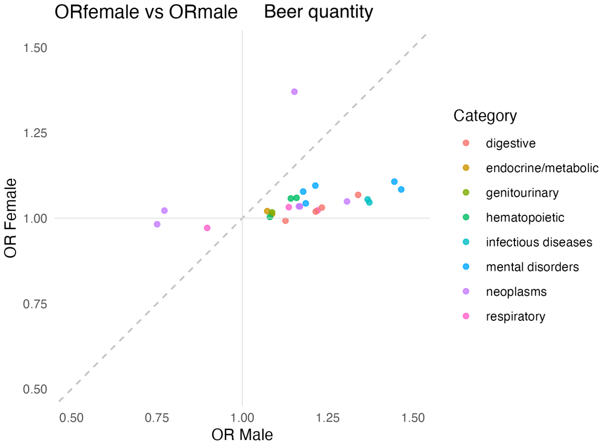
ORfemale vs ORmale Beer quantity
OR Female
OR Male
0.50
0.75
1.00
1.25
1.50
Category
digestive
endocrine/metabolic
genitourinary
hematopoietic
infectious diseases
mental disorders
neoplasms
respiratory

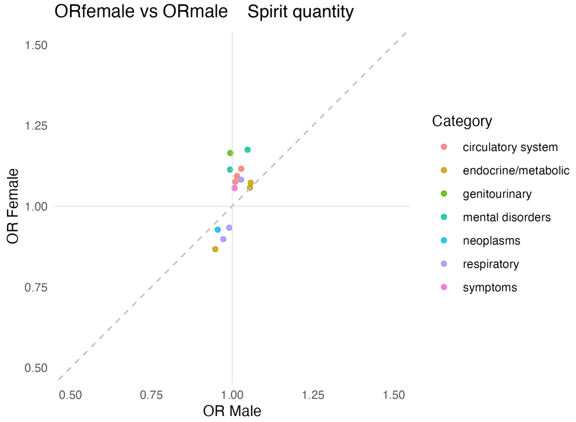
ORfemale vs ORmale Spirit quantity
OR Female
OR Male
0.50
0.75
1.00
1.25
1.50
Category
circulatory system
endocrine/metabolic
genitourinary
mental disorders
neoplasms
respiratory
symptoms

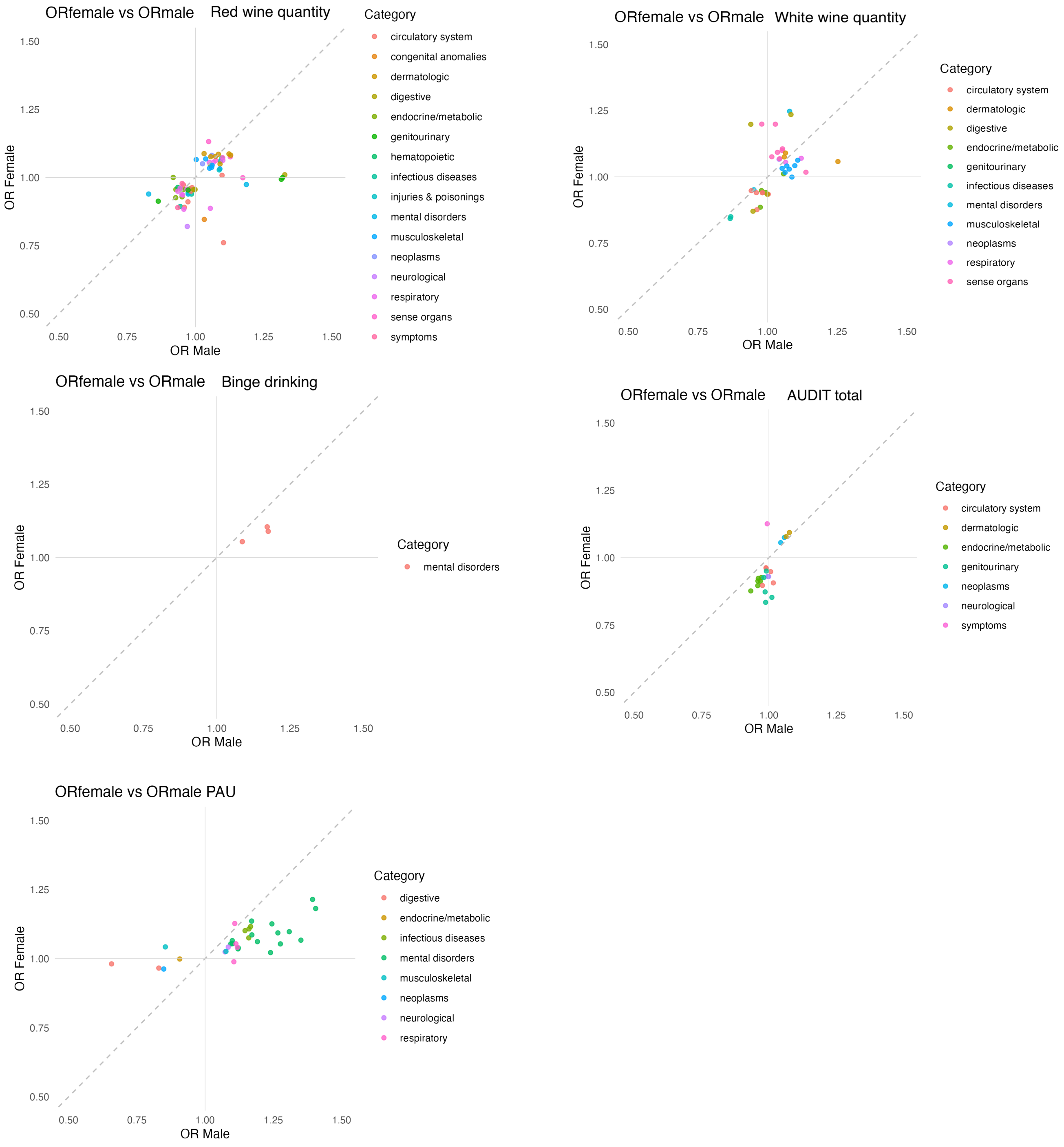

**Figure S6.** Comparison of female- and male-specific phenome-wide association study (**PheWAS**) results for each alcohol use phenotype. The y-axis shows odds ratios (**OR**) from female-specific polygenic scores (**PGS**) associations, and the x-axis shows OR from male-specific PGS associations for traits with significant sex-differentiated effects. The color of the dots corresponds to the trait category.

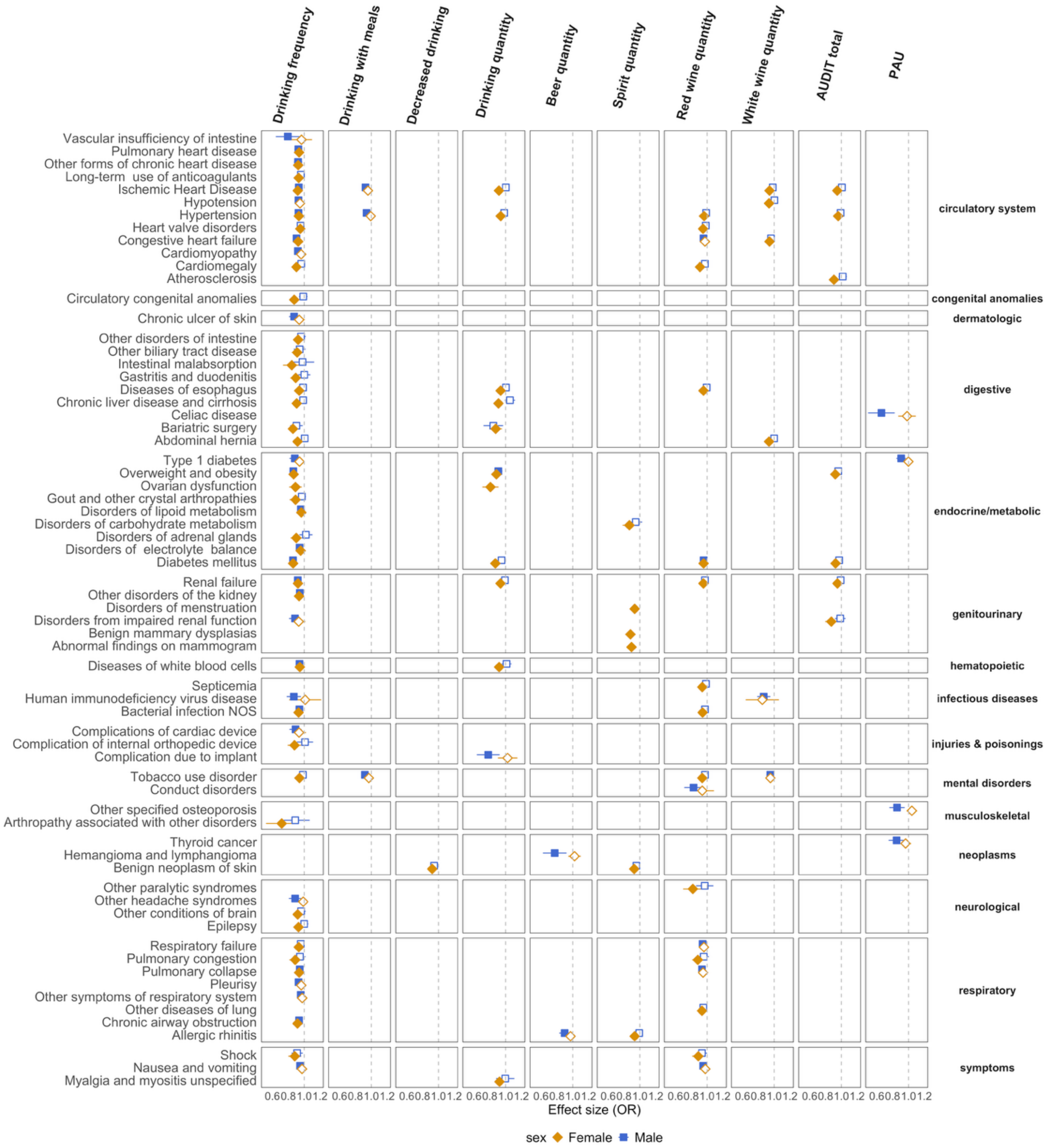

**Figure S7.** *Negative* associations between alcohol use phenotypes PGSs and medical conditions across disease categories from the PheWAS results. Results are displayed for female PGS in the female target sample (amber diamonds), and male PGS in the male target sample (blue squares) PGSs. OR are shown for selected phenotypes exhibiting negative associations. Filled diamonds/squares represent FDR-corrected significant associations, while empty ones indicate non-significant associations.
