## Supplementary Material for "Sex-Specific Genetic Architecture and Comorbidities of Alcohol Use Behaviors"

### METHODS

#### Data selection

Phenotypes were selected based on prior evidence of differences in between-sex genetic correlation ($r_g$ < 0.9; Savage et al., 2024). We additionally excluded phenotypes with non-significant or low $h^2_{SNP}$ in the sex-combined sample ($h^2_{SNP}$ < 0.03 and/or Z-score < 10) to ensure reliable results (Lee et al., 2018). The full list of phenotypes excluded based on these criteria is shown in **Figure S1**. Two phenotypes were excluded due to $r_g$ > 0.9, including *auditc_ln* (AUDIT-C score) and *pershistory* (self-report of ever having physical addiction to alcohol). Six phenotypes were excluded due to $h^2_{SNP}$ < 0.03, including *abuse* (Harmful/risky use), *advice* (received counseling from medical practitioner about alcohol use), *anyclin* (any clinically significant event related to alcohol misuse), *broad_aud* (broad alcohol use disorder definition), *increasedrink* (drinking has increased over the past 10 years) and *quantfwine* (quantity of fortified wine consumed per month). Lastly, *AUDIT-P* was excluded because it is a subset of the *PAU* sample (**Figure S1**). Further details on the excluded phenotypes can be found in the original publication (Savage et al., 2024).

#### AUDIT total score sex-stratified GWAS

We performed sex-combined and sex-stratified GWAS of AUDIT total score using new data from 72,444 female and 40,335 male research participants from 23andMe Research Institute. Participants provided informed consent and volunteered to participate in the research online, under a protocol approved by the external AAHRPP-accredited Salus IRB (https://www.versiticlinicaltrials.org/salusirb). Quality control, imputation, and genome-wide analysis were conducted by 23andMe as previously described (Sanchez-Roige et al., 2023). Association tests were conducted via linear regression under an additive model using a proprietary pipeline developed internally by 23andMe, controlling for age (inverse-normal transformed), sex, the top five principal components (**PC**) of genotype, and indicator variables for genotype platforms. The AUDIT (Saunders et al., 1993) was administered to assess alcohol use over the past year. AUDIT total score was created by taking the sum of items 1–10 for all participants, including those who endorsed currently never drinking alcohol (as they could still endorse past hazardous use on items 9 and 10). The score was log10-transformed to approximate a normal distribution.

We identified lead SNPs using the clumping function in PLINK v.2.0 (Chang et al., 2015) with an $r^2$ threshold of 0.1, a distance of 500 kb and the 1000 Genomes Project phase 3 as the LD reference panel. Lead SNPs located <1 Mb apart were merged into one locus.

### RESULTS

#### GWAS of AUDIT total scores

In the sex-combined sample, we identified 5 genome-wide significant independent loci ($P < 5 \times 10^{-8}$) associated with AUDIT total scores; two of which replicated prior alcohol GWAS (Sanchez-Roige et al., 2019; Zhou et al., 2020). In the female-stratified analysis (N = 72,444, **Table S1, Figure S3**), we found three genome-wide significant loci. The top hit (rs3114045, *P* = 7.40e-13) was located on chr4q23 between the *ADH1B* and *ADH1C* genes, replicating previous associations for several alcohol GWAS (e.g., AUDIT total score and the addiction risk factor GWAS (Hatoum et al., 2023; Sanchez-Roige et al., 2019). In the male-stratified analysis (N = 40,335, **Table S1, Figure S3**) we identified one genome-wide significant loci that replicated the top hit in the sex-combined sample (lead SNP: rs138423208, *P* = 4.80e-08), which was located on chr4q23 between the *ADH6* and *ADH1A* genes. We noted a nominally significant difference in $h^2_{SNP}$ estimates for AUDIT total score, which were higher in males than females (0.097 ± 0.014 vs. 0.068 ± 0.010, *p* = 0.026), despite the lower sample size in the male cohort (**Table S2**, **Figure S2**).

### REFERENCES SUPPLEMENTARY MATERIAL


Chang, C. C., Chow, C. C., Tellier, L. C., Vattikuti, S., Purcell, S. M., & Lee, J. J. (2015). Second-generation PLINK: Rising to the challenge of larger and richer datasets. *GigaScience*, *4*, 7. https://doi.org/10.1186/s13742-015-0047-8

Hatoum, A. S., Colbert, S. M. C., Johnson, E. C., Huggett, S. B., Deak, J. D., Pathak, G., Jennings, M. V., Paul, S. E., Karcher, N. R., Hansen, I., Baranger, D. A. A., Edwards, A., Grotzinger, A., Substance Use Disorder Working Group of the Psychiatric Genomics Consortium, Tucker-Drob, E. M., Kranzler, H. R., Davis, L. K., Sanchez-Roige, S., Polimanti, R., … Agrawal, A. (2023). Multivariate genome-wide association meta-analysis of over 1 million subjects identifies loci underlying multiple substance use disorders. *Nature. Mental Health*, *1*(3), 210–223. https://doi.org/10.1038/s44220-023-00034-y

Lee, J. J., McGue, M., Iacono, W. G., & Chow, C. C. (2018). The accuracy of LD Score regression as an estimator of confounding and genetic correlations in genome-wide association studies. *Genetic Epidemiology*, *42*(8), 783–795. https://doi.org/10.1002/gepi.22161

Sanchez-Roige, S., Jennings, M. V., Thorpe, H. H. A., Mallari, J. E., van der Werf, L. C., Bianchi, S. B., Huang, Y., Lee, C., Mallard, T. T., Barnes, S. A., Wu, J. Y., Barkley-Levenson, A. M., Boussaty, E. C., Snethlage, C. E., Schafer, D., Babic, Z., Winters, B. D., Watters, K. E., Biederer, T., … Palmer, A. A. (2023). CADM2 is implicated in impulsive personality and numerous other traits by genome- and phenome-wide association studies in humans and mice. *Translational Psychiatry*, *13*(1), 167. https://doi.org/10.1038/s41398-023-02453-y

Sanchez-Roige, S., Palmer, A. A., Fontanillas, P., Elson, S. L., 23andMe Research Team, the Substance Use Disorder Working Group of the Psychiatric Genomics Consortium, Adams, M. J., Howard, D. M., Edenberg, H. J., Davies, G., Crist, R. C., Deary, I. J., McIntosh, A. M., & Clarke, T.-K. (2019). Genome-Wide Association Study Meta-Analysis of the Alcohol Use Disorders Identification Test (AUDIT) in Two Population-Based Cohorts. *The American Journal of Psychiatry*, *176*(2), 107–118. https://doi.org/10.1176/appi.ajp.2018.18040369

Saunders, J. B., Aasland, O. G., Babor, T. F., de la Fuente, J. R., & Grant, M. (1993). Development of the Alcohol Use Disorders Identification Test (AUDIT): WHO Collaborative Project on Early Detection of Persons with Harmful Alcohol Consumption--II. *Addiction (Abingdon, England)*, *88*(6), 791–804. https://doi.org/10.1111/j.1360-0443.1993.tb02093.x

Savage, J. E., Barr, P. B., Phung, T., Lee, Y. H., Zhang, Y., McCutcheon, V. V., COGA Investigators, Ge, T., Smoller, J. W., Davis, L. K., Meyers, J., Porjesz, B., Posthuma, D., Mallard, T. T., & Sanchez-Roige, S. (2024). Genetic Heterogeneity Across Dimensions of Alcohol Use Behaviors. *The American Journal of Psychiatry*, *181*(11), 1006–1017. https://doi.org/10.1176/appi.ajp.20231055

Zhou, H., Sealock, J. M., Sanchez-Roige, S., Clarke, T.-K., Levey, D. F., Cheng, Z., Li, B., Polimanti, R., Kember, R. L., Smith, R. V., Thygesen, J. H., Morgan, M. Y., Atkinson, S. R., Thursz, M. R., Nyegaard, M., Mattheisen, M., Børglum, A. D., Johnson, E. C., Justice, A. C., … Gelernter, J. (2020). Genome-wide meta-analysis of problematic alcohol use in 435,563 individuals yields insights into biology and relationships with other traits. *Nature Neuroscience*, *23*(7), 809–818. https://doi.org/10.1038/s41593-020-0643-5
